# Consistent DNA methylation patterns enable accurate and interpretable cross-platform classification of central nervous system tumors

**DOI:** 10.1101/2025.10.07.25337348

**Authors:** Elaheh Moradi, Juuso Vuorinen, Tomi Hoikka, Anja Hartewig, Laura Helin, Alejandra Rodriguez-Martinez, Meeri Pekkarinen, Miina Vulli, Suvi Lehtipuro, Sinikka Ampuja, Artturi Mäkinen, Vidal Fey, Francesco Tabaro, Andries De Koker, Ruben Van Paemel, Bram De Wilde, Nico Callewaert, Milla E.L. Kuusisto, Hanna-Riikka Teppo, Outi Kuittinen, Hannu Haapasalo, Kristiina Nordfors, Matti Nykter, Joonas Haapasalo, Juha Kesseli, Kirsi J. Rautajoki

## Abstract

**Background:** DNA methylation–based classification has become an integral component of central nervous system (CNS) tumor diagnostics in neuro-oncology. However, current classifiers would benefit from improved interpretability, cross-platform generalizability, and scalability in routine clinical practice.

**Methods:** We developed a hybrid feature selection and machine-learning framework to derive compact, biologically relevant DNA methylation feature sets for CNS tumor classification. Variance-based filtering, intra- and inter-class consistency assessment, and elastic-net logistic regression were combined to identify informative CpG regions. A linear support vector machine (SVM) classifier was trained to distinguish methylation classes using microarray data and applied for sequencing data after imputing missing values. Selected tumor classes were differentiated with a few informative classifying features.

**Results:** The framework identified 1,003 informative genomic regions enriched for enhancer elements and neurodevelopmental pathways. In large external validation cohorts profiled by DNA methylation arrays (n = 1,993), the classifier achieved an accuracy of 0.96. The low-dimensional feature sets supported the investigation of diagnostically challenging cases and improved differentiation of histologically similar tumor entities, like embryonal tumors, using as few as two discriminative CpG features. Robust classification was preserved with bisulfite-equivalent targeted methylation sequencing and untargeted Nanopore-sequencing data. *MGMT* promoter methylation state and off-target read-derived genome-wide DNA copy number profiles provided supportive information.

**Conclusions:** Consistent DNA methylation patterns combined with SVM enable accurate and interpretable CNS tumor classification across sequencing platforms and provide a clinically scalable framework for next-generation neuro-oncology diagnostics.

**Key points:** Robust cross-platform CNS tumor classification using 163-1,003 CpGs’ methylation call Genome-wide DNA copy number profiles from off-target reads of targeted sequencing Specific tumor types can be accurately separated using just two CpG features

**Importance of the study:** This study reports a set of informative DNA methylation features for CNS tumor classification together with their DNA methylation patterns. It introduces an accurate machine learning classification approach for CNS tumors. By selecting 1,003 informative features through a hybrid feature selection, the model achieved high classification accuracy (0.96) using support vector machines. Strong performance is maintained even with 163 features. Our method performs robustly across sequencing platforms and sample types, with a possibility to obtain DNA copy number profiles and other supportive information. This flexibility, reduced complexity, cost-effectiveness, and low-input requirements for sequencing makes it a promising, scalable option for routine use in clinical neuro-oncology. By focusing on select genomic regions, the model enhances interpretability, improves diagnostic confidence, and allows separation between classes with only a few features. In short, our shared features and model increase explainability, providing avenues to support and complement existing neuro-oncology diagnostics.

## Introduction

CNS tumors constitute a large and heterogeneous group of entities ^1^, that are among the most diagnostically challenging neoplasias. Accurate classification is essential for clinical decision-making and treatment planning. Traditionally, histological methods have been considered the gold standard for diagnosing CNS tumors ^1^. Despite its demonstrated utility, histological diagnosis also has limitations such as inter- and intraobserver variability ^2^ as well as site-related discrepancies ^3^, leading to misdiagnosis and inconsistent classifications.

DNA methylation profiling can partly overcome histology-based diagnostic limitations ^1,4,5^. DNA methylation profiles represent stable and tumor-specific molecular patterns, making them informative for diagnostic purposes ^6–8^. A methylation-based random forest classifier developed by Capper *et al*. ^4^ (hereafter referred to as the Molecular Neuropathology (MNP) classifier) is widely used for integrated diagnosis in clinical settings ^4^. Integration of histological and DNA methylation-based assessments led to diagnostic changes in 12% of the cases in the seminal study (71% involving WHO grade modifications) ^4^ and in 14% and 10% of cases in subsequent implementation studies ^9,10^, with a similar proportion of WHO grade changes (71%) in the latter. The importance of methylome profiling is underscored by its integration as a diagnostic tool in the most recent WHO classification, also giving rise to new entities uncovered via DNA methylation analysis ^1,11^.

Methylation profiling with microarrays that measure up to 935,000 and currently use up to 10k CpG sites for classifying (EPIC v2.0 with MNP v12.8) is already in clinical use. However, dependence on this time-consuming, special instrumentation-dependent, and expensive workflow still limits its broader use in routine diagnostics. Additional factors, such as low tumor cell content and poor DNA quality, may lead to low confidence results ^4^. On top of the technical burden, most models function as black boxes, providing little insight into why a sample is assigned to a particular class or how close competing diagnoses are. CNS tumor classification using cell-free DNA (cfDNA) has already been recently demonstrated to be feasible using cerebrospinal fluid (CSF) and plasma ^12,13^. Unexpected or borderline classifications still remain difficult to interpret, slowing down the wider adoption of methylation-based diagnostics.

To address these limitations, we developed a compact and interpretable DNA methylation classifier for CNS tumors. Using a hybrid feature-selection strategy, we identified 1,003 CpG sites that carry a strong diagnostic signal and capture biologically meaningful differences between tumor types. With these features, a linear support vector machine (SVM) classifier achieves accuracy comparable to that of high-dimensional models while using only a fraction of the original input space. After imputing missing values, our feature set also performs well across tested sequencing approaches. Notably, the feature set can be compressed further to 163 CpGs with minimal loss of performance, opening the door to cost-efficient targeted panels.

## Materials and methods

### Processing of DNA methylation microarray data

To construct a dataset for feature selection, we downloaded IDAT files from the CNS tumor reference cohort established by Capper *et al*. (GSE90496) from the Gene Expression Omnibus (GEO). For feature selection using the multi-class consistency approach, we processed the files together with a set of 36 blood and 30 cfDNA samples (including samples from GSE122126, GSE109430, GSE77056, and GSE128654) to allow the expansion of cfDNA analysis in follow-up projects. We read the files into R using minfi ^14^, and filtered, batch-effect corrected, and normalized the data as described by Capper *et al*. ^4^ (Supplementary Methods). The preprocessed dataset, referred to as the reference cohort, contained 2,801 samples and 428,230 probes.

For building a validation cohort, we similarly downloaded and pre-processed the validation dataset of Capper et al. (with v11b class labels) (GSE109379) from GEO, which resulted in an additional 1104 samples and 428,799 probes. We then constructed another validation set from six cohorts (GSE147548, GSE125450, GSE198855, GSE156090, GSE193196, and GSE104210), resulting in 899 samples and 428,230 probes. The GSE337222 data set was processed similarly for validating the binary classification results. We determined methylation-based classes for samples without prior classification results available using the next-generation molecular neuropathology (MNP) v11b4 (version 3.3) platform (v11b labels) that was hosted by the German Cancer Research Center (DKFZ) and version v12.8 accessible via Epignostic (https://app.epignostix.com/).

### Feature selection and characterization

We refined the original feature set with a hybrid feature selection strategy combining filtering approaches and Elastic Net Logistic Regression (ENLR) implemented with glmnet package (version 4.1-7) (Supplementary Methods). For the filtering, we categorized tumor classes based on consistency and used intra-class and multi-class consistency criteria in parallel to achieve reduced feature sets of 4,109 and 1,826, respectively. Union of these, 4,976 features, was used in the feature selection with ENLR. We conducted 10 runs of 10-fold cross-validation (CV) to gain 100 models, extracted the absolute coefficient values (ACV) derived from these models, and ranked the features based on them. We established two criteria to select the best feature set, namely average ACV>0.001 across the 100 models and the top ten most important features (key features) per class, and used the union of both outputs as the selected feature set. Consequently, the number of features was reduced from 4,976 to 1,003. The stability of reduction was assessed by calculating mean pairwise Jaccard similarity index (JSI) and coefficient variability across 100 ENLR runs (Supplementary Methods). We created further reduced feature sets by using stricter cutoffs for average coefficients and key features, which yielded 533, 298 and 163 when using cutoffs 0.002 and four, 0.01 and four, 0.01 and two respectively.

The selected 1003 and 4976 features were characterized based on their location in the genome and by performing gene set enrichment analysis (GSEA) (Supplementary Methods).

### Classifier

After selecting the relevant features, we applied SVM ^15–17^ with a linear kernel, to obtain the final predicted values (Supplementary Methods). We used R package e1071 supporting multiclass classification with the one-vs-one approach and the decision values to assign class labels. To assign confidence scores to our predicted classes, we utilized the predicted probability scores obtained from the SVM. We categorized the predicted classes into two groups to assess their confidence levels by defining a threshold for the relative difference between the probabilities of the best and second-best matches. We denoted the predicted probability of the best choice as Prob_1_ and the difference between the first- and second-best choices as Diff_12_ and categorized the samples into higher or low confidence group 1 and 2 based on the confidence score (Diff_12_/Prob_1_>0.5 and Diff_12_/Prob_1_<0.5), respectively.

### Targeted DNA methylation sequencing

Fresh-frozen (FF) or formalin-fixed paraffin-embedded (FFPE) tissue samples (n=47) were collected from 45 patients who underwent surgery between 2007 and 2025 at Tampere University Hospital Supplementary methods). Additionally, three patients underwent surgery at Oulu University Hospital. After pathological review (Supplementary Methods), genomic DNA was extracted from FF tissue samples using a QIAamp DNA Mini kit (Qiagen) or QIAamp Fast DNA Tissue Kit (Qiagen), and from FFPE samples with GeneRead DNA FFPE kit (Qiagen), using manufacturer’s instructions.

Libraries were prepared using 10 ng, 45 - 112 ng, and 600 ng of fragmented gDNA as input using manufacturers and developers recommendations with minimal deviations with cell-free reduced representation bisulfite sequencing (cfRRBS), targeted enzymatic methylation sequencing (EM-seq) and SureSelect methods, respectively (Supplementary Methods). We analyzed data using custom in-house pipelines and imputed missing values with median beta value of 7 nearest samples from combined GSE90496 and GSE109379 cohorts (Supplementary Methods). Copy number profiles were estimated from EM-seq off-target reads with QDNAseq (v1.42.0) and focal CNAs from tumor to normal ratio of total count normalized coverages from reads of targeted sites (Supplementary Methods). Cost and turn-around-time (TAT) were estimated using publicly available information (Supplementary Methods)

### Clinically meaningful methylation class separation with few consistent features

We selected consistent informative features that were differentially methylated in the index methylation class vs. the references and analyzed them as described in Supplementary Methods. The performance was tested in reference and validation cohorts and an external GEO dataset (GSE337222) with the selected beta value cutoffs (≤ 0.3 or ≤ 0.5).

### Visualizations

tSNE visualization ^18^ was done as described in Supplementary Methods and other visualizations using ggplot2 [27], ComplexHeatmap [28], karyoploteR [29], and gridExtra packages.

## Results

### Feature selection resulted in 1003 informative genomic regions

We used a reference cohort ^4^ with 82 CNS tumor methylation classes and nine control classes (v11b labels) for feature selection (Supplementary Table S1). The filtering of 32,000 most variable features using parallel intra-class and multi-class consistency approaches yielded an union of 4976 features and subsequent ENLR produced a robust panel of 1003 features (Jaccard similarity index = 0.85, mean coefficient SD = 0.00068 across 100 CV runs) (Figure 1A,B; Supplementary Figure S1A; Supplementary Table S2).

**Figure 1.**
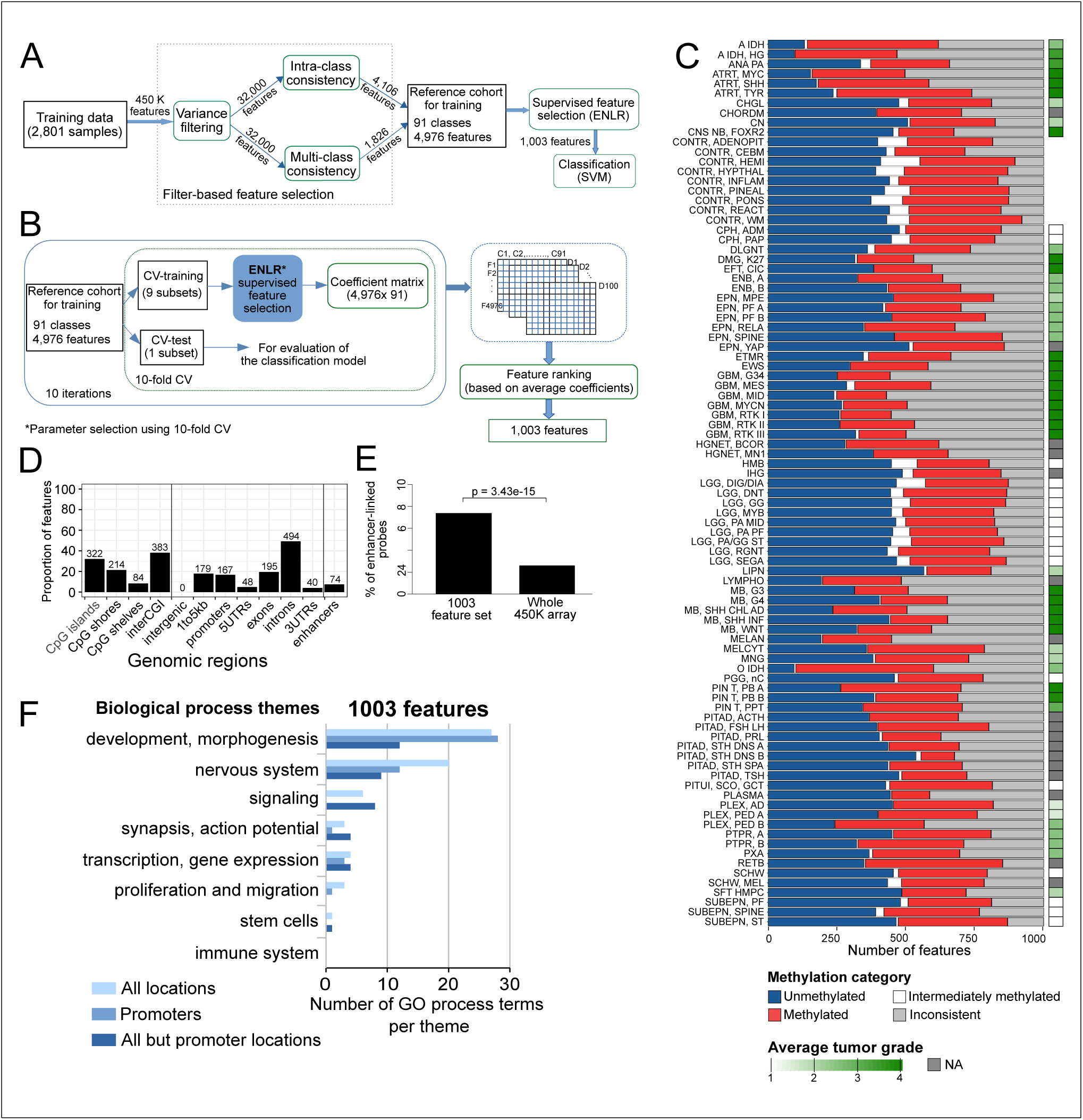
Characterization of the select 1003 features associate them with gene regulatory regions, nervous system, and development. (A) Schematic representation of the feature selection strategies. First, a variance filtering was applied followed by two parallel filtering-based strategies. Then, the results were pooled together. The resulting set of 4976 features was subjected to supervised feature selection by elastic net logistic regression (ENLR). Finally, classification was done using a support vector machine (SVM). (B) Schematic representation of the supervised ENLR-based feature selection. The reference cohort for training consisted of 4976 features measured from 2801 samples belonging to 91 tumor or control classes in total. A 10-fold cross-validation (CV) was used for feature selection by ENLR, and the process was repeated for 10 iterations, resulting in 1003 features. (C) Variable methylation patterns of the 1003 selected probes throughout DNA methylation classes and similarities among related tumor types. Distribution of features across methylation consistency categories (as defined in the multiclass consistency approach) for each tumor and normal methylation class is shown. Average tumor grade for each methylation class is shown on the right side of the bar plots. The tumor DNA methylation classes are ordered alphabetically. NA, no formal grading system. (D) The 1003 features were located mostly in introns, CpG islands, and interCGI regions. The percentage of features falling into different CpG island, gene, and enhancer locations for the 1003 feature set are shown. Absolute feature numbers for each genomic location are marked above the corresponding bar. (E) Features located in enhancers were over-represented in the selected 1003 feature set. The percentage of enhancer-linked features among the 1003 features as compared to the percentage in the Illumina Human Methylation 450k array after preprocessing is shown. Statistical significance was tested with Fisher’s exact test. (F) Development, morphogenesis and nervous system were the most common themes associated with genes located near to the CpGs contained in the 1003 features. Genes with CpGs in their promoter (proximal and distal) were more prone to be involved in development and morphogenesis than those with CpGs in other locations. The results from gene set enrichment analysis were filtered so that only Gene Ontology (GO) terms containing more than ten genes, with an enrichment p-value below 0.001 and with a fold enrichment above two were considered and grouped under Biological process themes.

We assessed the variability in consistent and inconsistent features between tumor classes by plotting the feature counts by methylation category for each class (Figure 1C), which showed related tumor classes to generally have similar consistent feature counts. Unmethylated features predominated in parts of pituitary adenomas and plasmacytomas. In contrast, methylated CpGs predominated in isocitrate dehydrogenase (IDH)-mutant diffuse gliomas and atypical teratoid/rhabdoid tumors (ATRTs). The vastly heterogeneous higher grade tumor classes showed a higher number of inconsistent probes in respect to the lower-grade tumors (Figure 1C).

Most of the 1003 selected features were intronic and located in CpG islands (CGI) and interCGI regions (Figure 1D). Features showed a 2.86-fold enrichment towards enhancers (p<0.0001, two-sided Fisher’s exact test) (Figure 1E). The 4976-feature set mirrored those characteristics (Supplementary Figure S1B). Both feature sets were uniformly distributed throughout the genome (Supplementary Figure S1C) within the limitations of the DNA methylation array measurements (Supplementary Figure S1D). Interestingly, the selected features were linked to genes associated with development, morphogenesis, and nervous system-related biological processes (Figure 1F, Supplementary Table S3), suggesting functional relevance.

The selected 1003 features were distributed into two main clusters, predominantly unmethylated (Cluster 1) or methylated (Cluster 2), based on their methylation profiles throughout the tumor and control classes (Figure 2). The number of selected features with a non-zero ENLR coefficient was rather evenly distributed across tumor methylation classes; the highest contributing feature counts were detected in some of the glioblastomas (GBMs) (Figure 2). As expected, tumor classes with highly similar consistent methylation profiles corresponded to methylation groups of the same or related tumor types, such IDH-mutant tumors or atypical teratoid rhabdoid tumors (AT/RTs) (Supplementary Figure S2, Supplementary Table S2). A rather unique DNA methylation profile was observed for cerebellar liponeurocytoma, while GBMs presented few differences in consistent features, not only among them, but also when compared with many other tumor classes (Supplementary Table S2).

**Figure 2.**
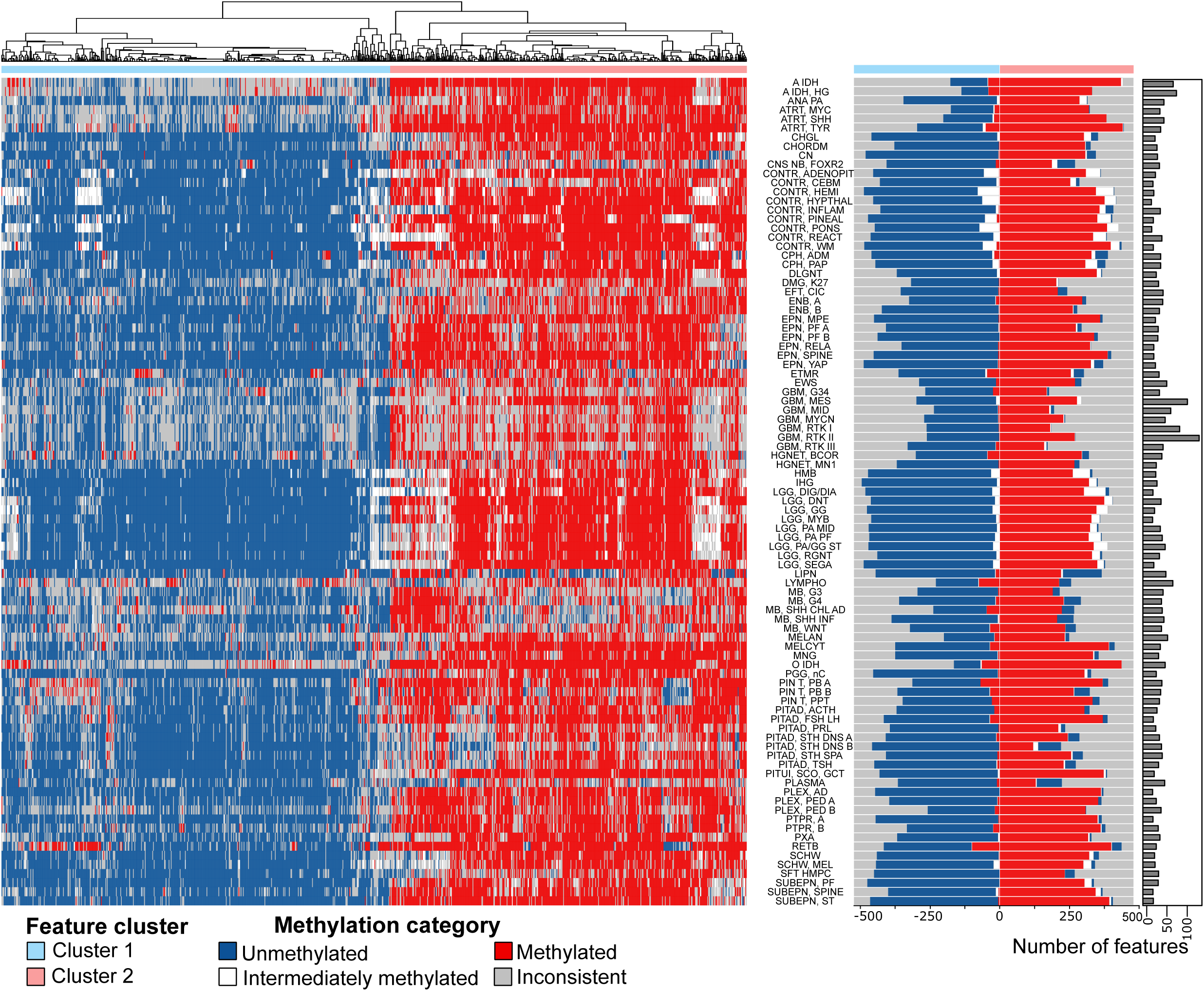
DNA methylation patterns of the select 1003 features across tumor methylation classes show a high degree of consistency. Inconsistent features are more predominant especially in diffuse gliomas and most embryonal tumors. Hierarchical clustering of the 1003 features according to their methylation categories identifies two distinct probe clusters, namely predominantly unmethylated probes (cluster 1, light blue) and predominantly methylated probes (cluster 2, light red). On the right side of the heatmap, the proportions of feature categories in the two identified feature clusters are visualized as stacked bar plots. Tumor classes with higher proportions of inconsistent features tend to include higher numbers of non-zero features in ENLR (gray bar plots on the right).

### High performance of the Support Vector Machine (SVM) classifier

Performance evaluation in the training phase used a 10-fold CV approach in the reference cohort (Figure 3A). The evaluation was performed at the methylation class level, and at diagnostic class level after grouping samples based on the 5th edition of the WHO’s CNS tumor classification (Supplementary Table S1). Using MNP DNA methylation class annotations as ground truth, overall and class-specific performance metrics were favorable in the reference cohort (Supplementary Figure S3A). These estimates should be interpreted as internal performance assessments because the initial variance- and consistency-based filtering step, despite being biologically motivated and not directly aiming to minimize classification error, was performed before cross-validation in the same cohort. Most misclassifications occurred within diagnostic classes (Supplementary Figure S3B, and Supplementary Table S1). At the diagnostic level, there were 17 misclassifications (0.61 %) compared to 64 (2.3 %) at the methylation class level, yielding good performance (Supplementary Figures S3B-D). To provide an independent assessment of classifier generalizability, performance was subsequently evaluated in an independent external validation cohort.

**Figure 3.**
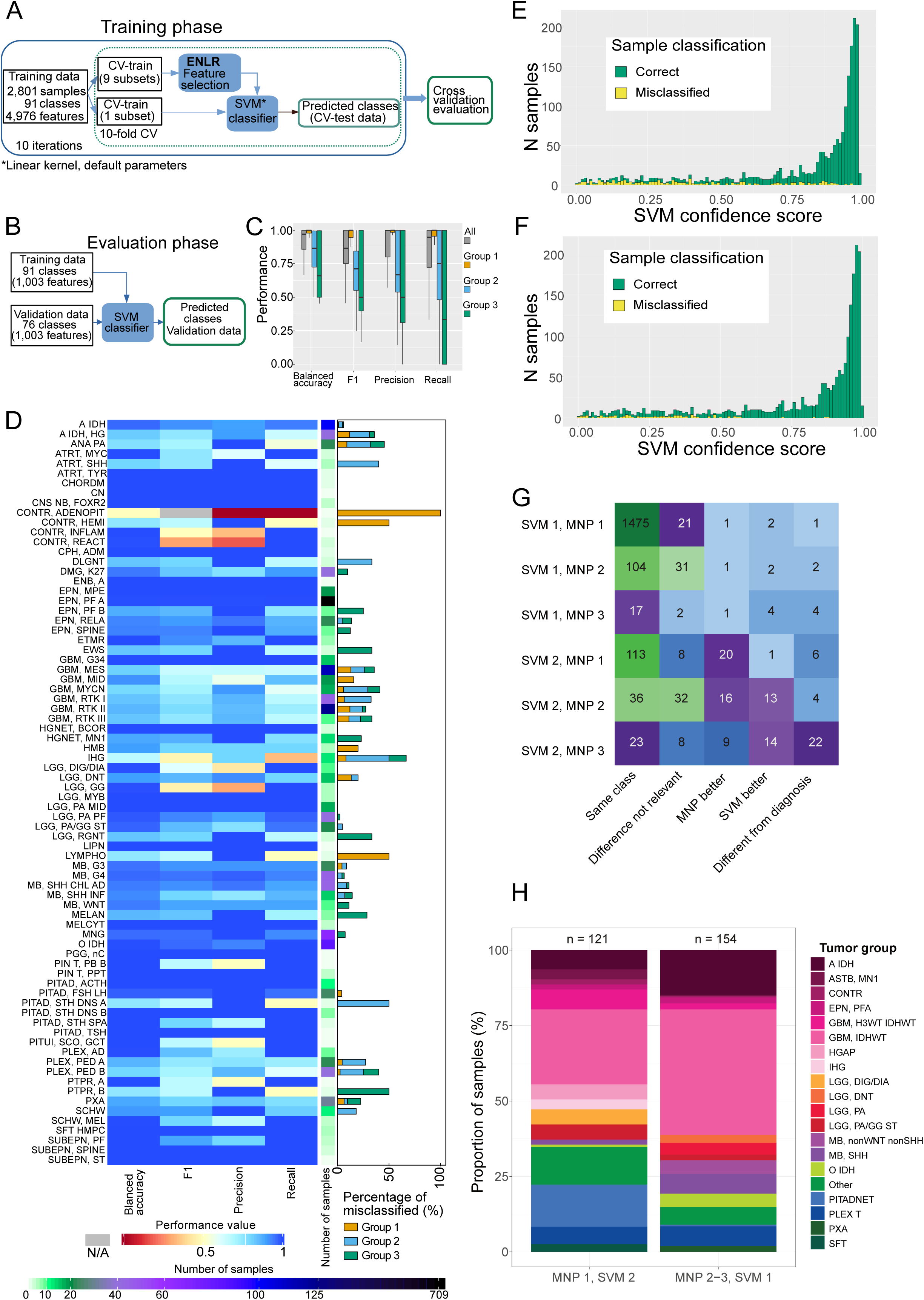
The performance of the SVM-based classifier is high in the DNA methylation array validation cohort (A) Schematic representation of the training phase of the SVM-based classifier. A 10 fold cross-validation (CV) was used for performance evaluation, and the whole process was repeated for 10 iterations. (B) Schematic representation of the evaluation phase of the SVM-based classifier. After selecting the optimal feature set, SVM was used for designing the final classifier. The final classifier was evaluated with an external validation data set. (C) Box plot showing the overall performance metrics (balanced accuracy, F1 score, precision, and recall) of SVM classification across tumor methylation classes based on MNP v11b class labels in the validation data. Results are shown for the whole dataset (All), and also after separating samples into three sample groups based on their calibrated MNP score. Group 1 includes samples with MNP score ≥0.84, Group 3 samples with score <0.5, and Group 2 rest of the samples. (D) DNA methylation classes with poorer performance include higher counts of misclassified low-confidence samples (samples in Group 2-3) or very few samples in general. Heatmap representation of the classification performance metrics across DNA methylation classes in the validation data when using MNP v11b classification as the ground truth. Total number of samples in the class, the percentage of misclassified samples, and their distribution to MNP confidence groups (Groups 1-3) are shown on the right. (E) Distribution of samples in the validation cohort based on the SVM confidence score. Classification accuracy in each sample was evaluated against MNP v11b classification. SVM confidence score of 0.5 was selected as a cutoff for high-confidence classification. (F) Distribution of samples in the validation cohort based on the SVM confidence score. Classification accuracy in each sample was evaluated so that irrelevant differences were ignored, and diagnosis-assisted assessment was used. (G) Nearly all of the samples (1658 of 1668, 99.4%) in SVM high confidence group (Group 1) are correctly classified. In the figure, the validation cohort samples are distributed into MNP (Groups 1-3, high confidence group: Group 1 with cutoff 0.84) and SVM (Groups 1-2) confidence groups (in rows), and the difference in MNP vs. SVM classification is shown in columns. For the majority of the samples, the DNA methylation class was the same (n=1768) or the difference between the classifications was not relevant for diagnostic purposes (n=102). Part of the samples were classified more accurately with MNP (n=48) or SVM (n=36). Samples with a different classification with MNP and SVM, and both classifications also deviating from the reported diagnosis (n=39), were predominantly detected in low-confidence classification groups. (H) Diagnostic tumor type distribution of samples which received high confidence classification (Group 1) only with MNP (on the left) or SVM (on the right) classification.

The primary evaluation of classifier performance was assessed using 1003 features with an external validation set of 1993 samples representing 72 tumors and four control methylation classes (Figure 3B and Supplementary Table S4). The validation dataset was divided into three groups based on the calibrated MNP Classifier scores: Group 1 (score ≥0.84, n=1648), Group 2 (0.5 ≤ score < 0.84, n= 241), and Group 3 (score <0.5, n= 104) (Supplementary Table S4). A calibrated score cutoff of 0.84 for group 1 has been reported to provide the best balance between sensitivity and specificity, and is also used in clinical settings ^10,19^. When the ≥0.90 was used as group 1 cutoff, as recommended by the original publication ^4^, 1589 of samples fall within this category.

We first assessed performance metrics (balanced accuracy, F1 score, precision, recall) by comparing SVM results to MNP classifications in the validation set (Supplementary Table S4). As expected, all the performance metrics were highest in group 1 (Figure 3C, Supplementary Figure S3E-I). Performance declined and variation increased with lower calibrated scores irrespective of used cutoff (0.84 or 0.9) (Figure 3C, Supplementary Figure S3E-F). DNA methylation class-specific performance metrics varied significantly between tumor types. GBMs, infantile hemispheric gliomas, IDH-mutant astrocytomas, pediatric plexus tumors (PLEX PED), and anaplastic pilocytic astrocytomas yielded the highest number of samples with discrepant classifications (Figure 3D). Again, most discrepancies occurred within the same diagnostic class (Supplementary Figure S3J-K). The use of selected features improved overall accuracy in the validation cohort, which was higher (0.888) when compared to randomly selected 1003 variable feature sets (mean 0.469, range: 0.463–0.475).

SVM concordance with MNP classification was very high among samples with high MNP confidence (96.42% accuracy; group 1, score ≥0.84), but decreased progressively with lower MNP confidence (Figure 3C, Supplementary Figure S3E–I). This raised concerns about using low-confidence MNP classifications as ground truth. We therefore assessed discrepant SVM and MNP classifications against the final integrated diagnosis (Supplementary Table S4), considering differences irrelevant when both classes belonged to the same tumor type and the difference did not affect treatment prediction or prognosis. This included age- or location-related subclasses and different GBM subtypes, which can coexist within the same tumor bulk ^20^. These assessments were termed diagnosis-assisted classifications. Among the 102 (3.6%) samples with irrelevant classification differences, both methods were considered consistent with the tumor diagnosis. MNP provided a more accurate classification than SVM in 48 (1.7%) cases, whereas SVM was more accurate in 36 (1.3%); in 39 (1.4%) cases, neither method matched the diagnosis. Applying the diagnosis-assisted criteria yielded an overall SVM accuracy of 0.957, and it was used in subsequent analyses.

To evaluate the confidence of SVM classification, the metric based on SVM probability and the difference from the second-best hit (hereafter referred to as confidence score) (Figure 3E-F) was used. Receiver Operating Characteristic (ROC) analysis using the external validation cohort and correct versus incorrect classifications as the outcome variable show that the confidence score discriminates well between correct and incorrect classifications (AUC = 0.92), and the optimal threshold determined using Youden’s index was 0.57 (sensitivity 0.85, specificity 0.95). Increasing the confidence threshold improved classification accuracy within the high-confidence group, with calibration analysis (Supplementary Figure S4) showing 100% accuracy among samples with confidence scores >0.9 (n = 1,092, 54.8% of validation cohort samples) but reduced the number of retained high-confidence samples (Supplementary Table S4). Confidence score ≥0.5 was selected as a criteria for high-confidence classification (called as Group 1). In the SVM group 1, 10 of the 1668 samples were misclassified (accuracy 0.996), three of which were correctly classified by MNP (Figure 3G, Supplementary Table S4).

In the SVM low-confidence group 2, 77 of the 325 samples were misclassified (accuracy 0.763) (Supplementary Table S4). These included 45 samples that were correctly classified only with MNP (20 of which were in MNP group 1). Recurrent SVM misclassifications in SVM group 2 included GBM MES tumors classified as CONTROL INFLAM or CONTROL REACT (n=11) and IDH-wild-type GBMs (n=16) classified as low-grade gliomas (LGGs) characterized by *BRAF* and other MAPK alterations (PXA and LGG GG, PA/GG ST, and DIG/DIA). Furthermore, one HMB, one DMG K27, and five PLEX PED A/B samples were classified as CONTR ADENOPIT in SVM Group 2.

SVM classification was found to be more accurate than that of MNP for 36 tumors (Figure 3G, Supplementary Table S4). All samples classified as IDH-mutant astrocytomas by MNP and as oligodendroglioma by SVM (n=6, two of which were in SVM group 1) were IDH-mutant oligodendrogliomas based on the final integrated diagnosis. Furthermore, the SVM classified correctly 10 GBMs and 11 LGG DIG/DIA samples misclassified by MNP. Most of the samples classified with high-confidence only with SVM (n=154, 7.7% of the validation cohort) were IDH-wild-type GBMs (42%) or IDH-mutant astrocytomas (15%) (Figure 3H, Supplementary Table S4).

When SVM classification performance was evaluated using MNP v12.8 class labels, same training cohort, and same set of 1003 features, the SVM v12.8 classification showed an accuracy of 0.826 (95% confidence interval (CI95) 0.804—0.845) in the validation microarray cohort (n=1344, a subset of the v11b validation set), and 0.934 (CI95 0.917—0.948) and 0.468 (CI95 0.413—0.523) in high and low confidence groups, respectively. Thirtythree tumors representing 19 classes present in the validation cohort but absent in the training set were removed from the analysis.

### Robust classification with DNA methylation sequencing across platforms

To demonstrate the utility of our selected DNA methylation features, we run SVM classification to different DNA methylation-sequencing samples, derived with targeted bisulfite-equivalent or Nanopore sequencing. We generated targeted EM-seq data from a cohort of 47 tumor samples (Supplementary Table S5) that were enriched using IDT xGen v2 (n=17) or xGen v3 (n=20) Hybridization Capture Core Reagents. The median feature coverage (i.e. the proportion of CpGs with at least 10 read coverage) was 62.7% (IQR 56.1—97.6%), 97.9% (IQR 97.5—98.1%), and 59.8% (IQR 49.8—62.5%) for whole cohort, batch 1, and 2, respectively (Supplementary Figure S5). Missing values were imputed using K-nearest neighbors imputation (Supplementary Table S6), which performed similarly well for microarray and EM-seq data (Supplementary Figure S6).

T-distributed stochastic neighbor embedding (t-SNE) analysis showed that most samples (except S13, S26, S39, S47, S48, S52, and S55) clustered with tumors of the corresponding diagnosis (Figure 4A). The SVM v11b classifier confirmed the histological diagnosis in 41/47 samples (accuracy 0.872, CI95 0.748—0.94) (Figure 4B), with all misclassified samples showing unexpected t-SNE localization (Figure 4A). S26 was correctly classified with SVM. Three of the misclassified tumors (S13, S52, S55) showed poor SVM confidence score (<0.5), improving the accuracy to 0.925 (CI95 0.801—0.974) among the high confidence classifications (Figure 4B, Supplementary Table S7). Two other misclassified samples, S39 (schwannoma classified as CONTR, HEMI) and S48 (IDH-wildtype diffuse astrocytoma which two years later relapsed as IDH-wildtype GBM, classified as LGG, PA/GG ST) had low estimated tumor contents (40% and 10%, respectively). Furthermore, S47 (ependymoma GII) was classified as ganglioglioma (LGG GG; confidence score 0.588, tumor fraction 80%) and showed extensive, complement-like calcification, which is atypical for ependymomas. Histopathological diagnosis was challenging due to limited ependymoma features (mainly focal perivascular pseudorosette-like growth pattern). The tumor could histologically and based on marker staining represent low-grade glioma but lacked convincing ganglioglioma features.

**Figure 4.**
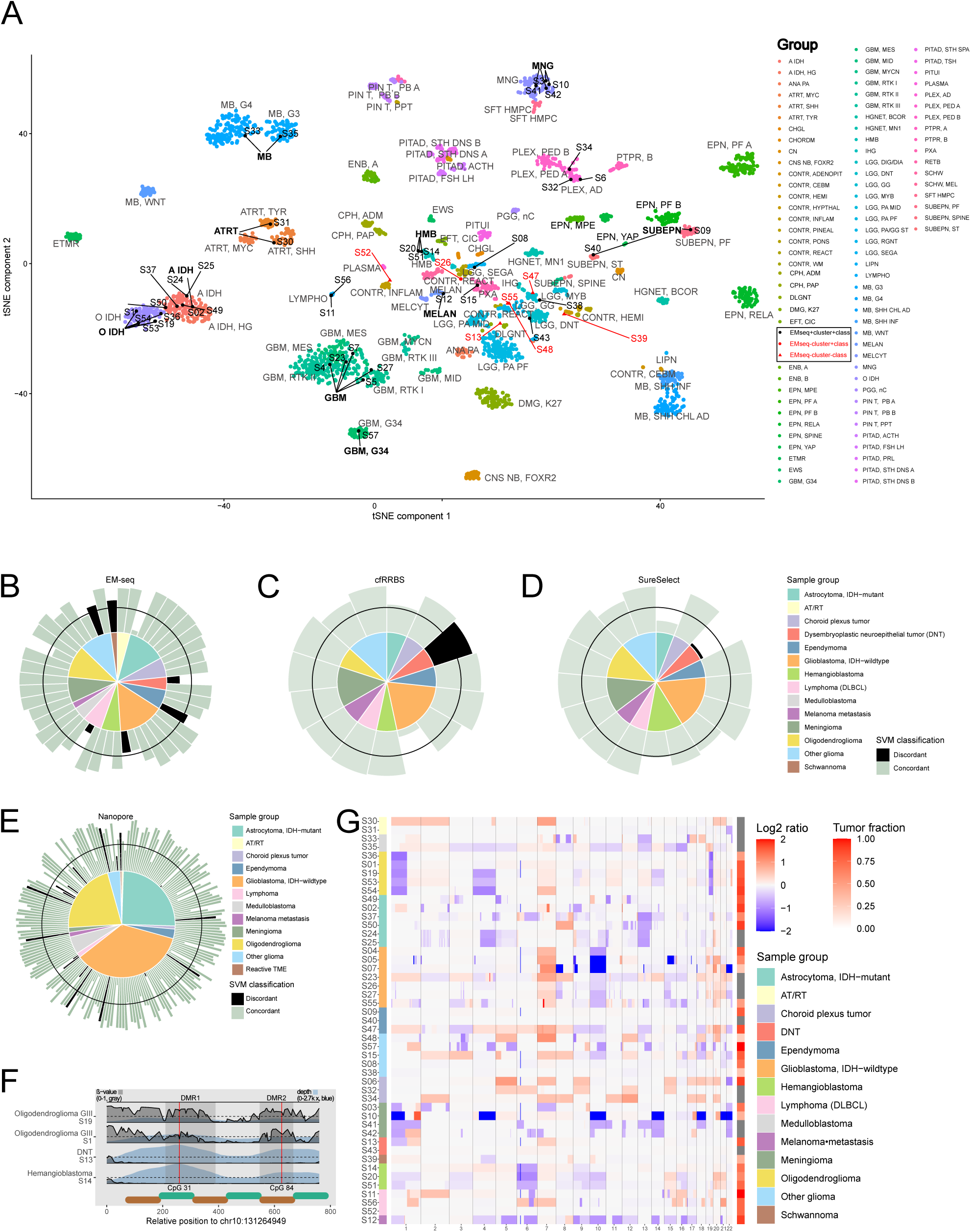
Targeted DNA methylation sequencing provides robust tumor classification, *MGMT* promoter methylation information, and genome-wide DNA copy number profiles. (A) t-SNE visualization of the reference cohort from Capper et al. (n=2,801), profiled using methylation arrays, together with our in-house cohort (n=47), profiled by targeted EM-seq. The analysis was based on 1,003 features, with missing values imputed in the EM-seq data. Most EM-seq samples (S1-S57) were positioned within or close to DNA methylation clusters corresponding to their tumor classes (shown in black), consistent with the clustering observed by Capper et al. Seven samples (S13, S26, S39, S47, S48, S52, and S55; red) showed discordant positioning. EM-seq samples with discordant classification are marked with triangles. Individual reference samples are colour-coded according to their methylation class. (B-E) Robust SVM-based v11b classification of sequenced tumor samples after imputing the missing values in EM-seq (n=47) (B), Sureselect (n=17) (C), cfRRBS (n=15) (D), and Nanopore (n=208) (E) sequencing cohorts. Samples are grouped based on their tumor type. For each sample, the classification confidence score (bar height) and concordance with the tumor diagnosis (green, concordant; black, discordant) are shown. Confidence score cutoff 0.5 is marked with a black line (circle) in each of these sunburst plots. (F) MGMT promoter methylation analysis of EM-seq data showed the expected methylated pattern in oligodendrogliomas compared with tumors in which MGMT promoter methylation is not typical, such as dysembryoplastic neuroepithelial tumor (DNT) and hemangioblastoma. Grey shading indicates beta values (0–1), and blue shading sequencing depth (0–2,728x). The dashed grey line indicates 0.3 beta value, a commonly used threshold for a methylated MGMT promoter. Brown and green bars indicate EM-seq target-enrichment probes across the MGMT promoter region relative to genomic coordinates (chr10:131,264,949). The locations of diagnostically relevant Differentially Methylated Region 1 (DMR1), DMR2, and commonly utilized microarray probes are highlighted. (G) DNA copy number alteration profiles obtained from off-target reads in the EM-seq data showcases typical patterns for the respective tumor types. Each row represents an individual sample. Colour intensity indicates tumor fraction-adjusted log2 ratios derived from normalized coverages relative to the diploid copy number, with the colour scale capped at −2 and 2. The bar on the left side of the plot depicts the tumor groups, whereas the bar on the right shows sample-wise estimated tumor fractions. Tumor fraction adjustment was not applied to samples without an estimated tumor fraction, marked in gray.

The v12.8 classifier produced largely similar results (Supplementary Table S7). Notably, S13 (dysembryoplastic neuroepithelial tumor, DNT) was classified with a higher confidence score (0.647 vs. 0.270 with v11b) as pilocytic astrocytoma (PA_INF).

Comparison of the 742 DNT-consistent CpGs covered in S13 showed discordant beta values for 104 (14.0%, including one imputed value), compared with a mean of 9.6% (83/868, range 73-94 features) in eight reference DNT microarray validation cohort samples, suggesting methylation heterogeneity. Neuropathological re-evaluation retained the DNT diagnosis but considered it uncertain. Although the tumor showed several typical DNT features, including multinodularity, oligodendroglial-like cells in the cell nodules, low proliferation, and columnar architecture, it lacked neuronal dysplasia, floating neurons, and a mucoid matrix and showed atypical increased lymphocyte counts in some regions, which may have influenced its methylation profile. Of note, distinguishing DNT from ganglioglioma and pilocytic astrocytoma can be challenging in the absence of typical separating features ^21,22^, and gross-total resection is a preferred treatment for all these tumors.

To further evaluate cross-platform applicability of our approach, we generated sequencing data with SureSelect hybrid capture assay and a method called cell-free reduced representation bisulfite sequencing (cfRRBS) ^23^ from 17 and 15 EM-seq batch 1 tissue samples, respectively. The data covered 602—678 and 160—194 of our features with at least 10 read coverage, respectively (Supplementary Table S6, Supplementary Figure S7A-D). Both cohorts included one misclassification (DNT, S13) (Supplementary Table S7). Furthermore, a publicly available untargeted Nanopore sequencing cohort of 208 measurements from 174 tumors and one control sample with a median of 730.5 features covered (range 100—1003) with at least 1 read coverage, was analyzed (Supplementary Figure S7E-F). In the Nanopore cohort, the predicted classes aligned with the original diagnosis with 0.904 accuracy (CI95 0.856—0.937) which increased to 0.954 (CI95 0.913—0.977) with high confidence classifications (confidence score >0.5) (Supplementary Table S7). Samples misclassified with a high confidence included two GBMs and a melanoma metastasis (classified as CONTROL INFLAM), a GBM (A IDH HG), a reactive tumor microenvironment (TME) sample (GBM MES), a pilocytic astrocytoma (subependymoma, SUBEPN PF), an IDH-mutant astrocytoma (LGG DNT), and an IDH-mutant oligodendroglioma (A IDH). Also in low-confidence classifications, three oligodendrogliomas were classified as IDH-mutant astrocytomas (A IDH class). Subpopulation of Nanopore samples (n=43) had corresponding microarray-based v11 classification data, showing Nanopore SVM classification accuracy of 0.930 (CI95 0.814—0.976) (Supplementary Table S7). The three misclassifications included two replicates of a CNS lymphoma classified with low confidence (0.125—0.377) as CPH PAP and the previously mentioned IDH-mutant astrocytoma (classified with a high confidence as LGG DNT), whose astrocytoma diagnosis was confirmed with microarray data. Other inconsistent classifications were between adult GBM subgroups or A IDH HG tumors classified as A IDH, which were not considered relevant.

To summarize, an accuracy of 0.872 (CI95 0.748—0.94) and 0.904 (CI95 0.856—0.937) was obtained with the EM-seq and Nanopore-seq-based v11b classification, respectively. Furthermore, the Sureselect and cfRRBS method-derived libraries showed similar performance (accuracy of 0.941 (CI95 0.73—0.99) and 0.933 (CI95 0.702—0.990), respectively) despite a significantly lower number of features covered in the data. Generally, the feature capture performance, coverage gained, cost and TAT favored targeted EM-seq with IDT XGen v2 reagents over the other short-read sequencing workflows (Supplementary Methods, Supplementary Figures S5 and S7).

### EM-seq provides supportive information about MGMT promoter methylation and genome-wide DNA copy-number profiles

The *MGMT* promoter region, as well as selected genomic loci harboring recurrent, diagnostically informative alterations, such as the *CDKN2A* gene, are covered by our EM-seq panel. *MGMT* promoter methylation analysis of the EM-seq data showed typical methylated patterns for all oligodendrogliomas (Figure 4F, Supplementary Table S8 and Supplementary Figure S8) ^24,25^.

The genome-wide DNA copy number alteration (CNA) patterns of the tumors, derived from off-target reads, show characteristic features such as chromosome 7 gain and 10 loss in all GBMs, co-deletion of chromosome 1p and 19q in all oligodendrogliomas, and, in all meningiomas, chromosome 22q deletion alongside other CNAs frequently observed in high-grade meningiomas (Figure 4G, Supplementary Figure S9) ^26^. The CNAs matched those detected from whole-genome sequencing of respective samples (S23-27) (Supplementary Figure S10). Targeted site CNA analysis revealed *CDKN2A* deletions in a subset of samples (Supplementary Figure S11).

### Accurate CNS tumor classification from a reduced numbers of features

SVM classification performance was also evaluated by including only a subset of 1003 features into the model. First, the 17 samples shared by enzymatic (EM-seq) or bisulfite (Sureselect, cfRRBS) sequencing cohorts were used. EM-seq, SureSelect methylation sequencing, and cfRRBS covered 986 (98%), 615 (61%) and 325 (32%) of the selected 1003 features, respectively, when 200bp distance cutoff to the index CpG and average methylation in ±50 bp region from the closest covered CpG was used (Supplementary Table S9 and Supplementary Figures S12).

After retraining an SVM classifier to each dataset, a good performance was observed. Again, all these samples were correctly classified, except S13 (Figure 5A), also misclassified using imputing (Figure 4B-D). SureSelect-based classification positioned S13 into a correct tumor class (DNT), albeit with a low confidence score (0.202) (Supplementary Table S9). Thus, accurate classification was achieved with DNA methylation sequencing using fewer than the original 1,003 features Figure 5A-B). Furthermore, performance dropped only marginally when we reduced features via ENLR-based selection to 533, 298, or 163 in the DNA microarray data (Figure 5C-D, Supplementary Table S10). SVM confidence score distribution remained largely similar with reduced feature counts, showing high confidence scores for most of the samples (Supplementary Figure S13). Classification accuracies upon a diagnosis-assisted assessment (as for the 1003 set above) remained high: 0.957, 0.955, and 0.947 for 533, 298, and 163 features, respectively (Supplementary Table S10), and the performance did not drastically drop in any separate DNA methylation class, either (Figure 5E). The robustness is supported by the measured DNA methylation values of 163 and 298 features in the reference and validation cohorts, with consistent DNA methylation levels within sample classes and clear differences between them (Supplementary Figure S14 and S15).

**Figure 5.**
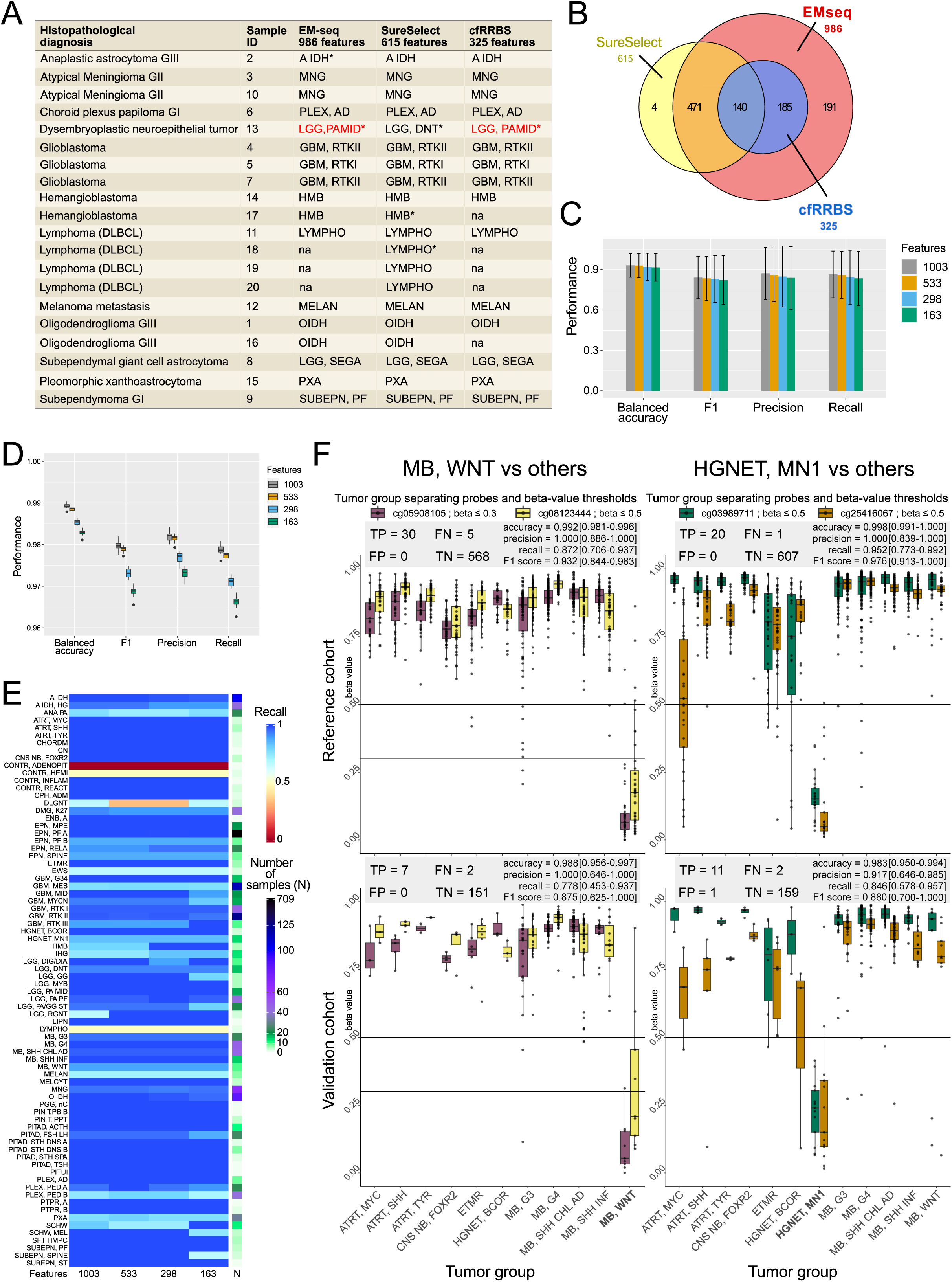
Tumor classification is feasible with even more reduced numbers of features. (A) Summary of SVM classifier labeling when applied to three different DNA methylation sequencing data that cover various amounts (986, 615, 325) of selected informative features. Similar high-precision classification is obtained even with just 325 features. All tumors were classified correctly with SureSelect data, although the SVM confidence score of sample 13 was poor with all the approaches. *: SVM confidence score <0.5, na: not analyzed (B) Only a moderate number of features are measured with all three targeted sequencing approaches. Venn diagram showing the overlap between the set of selected 1003 informative features (990 of which are covered by at least one method) and different reduced sets measured with DNA methylation sequencing approaches. Regions in which the maximum distance to the closest measured CpG was 200 bp from the probe coordinate were included in the analysis. (C) Performance metrics of the SVM v11b classifier in the microarray validation cohort are only slightly decreased when the number of features is reduced by adjusting the ENLR parameters. (D) Reduction of feature numbers by adjusting the cutoffs for ENLR-based feature selection has a moderate impact on the performance parameters of the SVM v11b classifier in the training phase. Data represents performance in the microarray reference cohort. (E) Classification performance remains similar across DNA methylation classes when the number of features was reduced from 1003 to 163. Heatmap representing the classification recall rate for each v11b class when irrelevant differences were ignored and diagnosis-assisted assessment was used. Samples were distributed to the classes based on MNP class labels. (F) Two diagnostically relevant classes named “Medulloblastoma, WNT-activated” (MB, WNT) and “Astroblastoma, MN1-altered” (represented by HGNET, MN1), can be separated with near perfect precision from the reference embryonal tumor classes using only two features for each comparison. Distinguishing the MB, WNT using probes cg05908105 (beta ≤ 0.3, or beta ≤ 0.5) and cg08123444 (beta ≤ 0.5) resulted in accuracy of 0.992 [0.981-0.996] and 0.988 [0.956-0.997] for the reference and validation cohorts, respectively. Probes cg03989711 and cg25416067 (beta ≤ 0.5 for both) reached accuracy of 0.998 [0.991-1.000] and 0.983 [0.950-0.994] in separating HGNET, MN1 cases from embryonal tumors in the reference and validation cohorts, respectively. Ninety-five percent confidence intervals (in square-brackets) for accuracy, precision, and recall were calculated using Wilson score intervals, and confidence intervals for the F1 score were estimated by nonparametric bootstrap resampling (100,000 iterations). TP: True positive, FN: False negative, FP: False positive, TN: True negative.

### Consistent features allow accurate tumor type separation

To take full advantage of our consistent feature set, we investigated whether one selected tumor class can be distinguished from other histologically similar tumor types in a clinically relevant manner using only a handful of DNA methylation features. This would allow differential diagnostics with a readily accessible methodology, such as methylation-specific quantitative PCR assays. In this analysis, samples were allocated into DNA methylation classes based on MNP classification. We also tried to separate anaplastic pilocytic astrocytoma from GBMs but could not find any suitable probes to that (Supplementary Table S2).

MB, WNT medulloblastomas are more treatment responsive than other medulloblastomas and embryonal tumors, and, thus, they are treated less aggressively. Their rapid differential diagnosis opens up opportunities for early start of adjuvant therapies. Comparison of MB, WNT against other embryonal tumors resulted in the identification of two probes, cg05908105 and cg08123444, which are methylated in other embryonal tumor types and show an unmethylated pattern in the index tumor type (Figure 5F, Supplementary Table S2). Both features were originally selected with the intra-class consistency approach requiring ≥ 90% consistency within the class. The CpGs measured by these probes are located in ENCODE cis regulatory elements (CREs) with enhancer marker H3K27ac positivity ^27^. The cg05908105 probe targets a CpG in the 3rd intron of NET1. The cg08123444 site is not annotated to any gene and it is located in the intergenic region upstream of *YWHAQ*, *TAF1B*, and *ADAM17* genes. Interestingly, both NET1 and ADAM17 have been associated with higher WNT activity ^28,29^.

In the reference cohort, we were able to separate MB, WNT tumors (n=39) from other embryonal tumors (n=568) based on these probes’ methylation levels that were below the selected thresholds (beta ≤ 0.3 for cg05908105 and beta ≤ 0.5 for cg08123444) with the accuracy of 0.992 (F1 score: 0.932, precision: 1.000, recall: 0.872) (Figure 5F). Performance did not change when a cutoff of beta ≤ 0.5 was used for both probes. Similar high performance with no false positives was observed in the external validation cohort comprising 9 MB, WNT cases and 151 other embryonal tumors (accuracy: 0.988, F1 score: 0.875, precision: 1.000, recall: 0.778) and in an independent cohort containing 52 MB, WNT cases and 253 other MBs (accuracy: 0.974, F1 score: 0.917, precision: 1.000, recall: 0.846) (Supplementary Figure S16). Out of two false negative samples in the first validation cohort, one had been classified with high confidence and one with low (MNP classifier score 0.496). Beta values of the probe cg08123444 were above the threshold in both, albeit lower beta value (0.60) was detected in the sample with high-confidence classification.

The differential diagnosis of astroblastoma, MN1-altered glioma (represented by HGNET, MN1 DNA methylation class) in respect to embryonal tumors can be challenging. Similarly to the above analysis, the comparison of HGNET, MN1 cases against all the embryonal tumors, including the MB WNT, resulted in the identification of two probes, cg03989711 and cg25416067, which were predominantly unmethylated in index tumor group and methylated in embryonal tumors (Figure 5F). Both features were originally selected with the intra-class consistency approach requiring ≥ 90% consistency within the class. The CpGs measured by these probes are located in ENCODE CREs with enhancer marker H3K27ac positivity ^27^. The CpG measured by cg03989711 is located in the 5th intron of *B3GNTL1*, but it is also upstream of the *METRNL* gene, whereas the cg25416067 target CpG is within the 1st exon or 2nd intron of *SEPTIN9*, depending on the transcript variant. Both *METRNL* and *SEPTIN9* have been linked to neural differentiation ^30,31^.

Requiring lack of methylation from both probes for a sample to be classified as HGNET, MN1 (beta value < 0.5), HGNET, MN1 tumors (n=21) were accurately separated from embryonal tumors (n=607) in the reference cohort with the accuracy of 0.998 (F1 score: 0.976, precision: 1.000, recall: 0.952). Again, high separation performance was also detected in the validation cohort comprising 13 HGNET, MN1 cases and 160 embryonal tumors (accuracy: 0.983, F1 score: 0.880, precision: 0.917, recall 0.846). The reference cohort contained no false positives. However, one false positive case was detected in the validation cohort. The MNP classified the case to MB, WNT group with a very low classification score (0.122), and it had been diagnosed as CNS Embryonal tumor, NOS (Supplementary Table S4), reflecting some uncertainty in the diagnosis. MNP classified two false negative samples in the validation cohort as HGNET, MN1 with low classification scores (0.046 and 0.447). In the pathology evaluation, the first case had been diagnosed as Embryonal tumor with multilayered rosettes, C19MC-altered (ETMR) and the other as anaplastic ependymoma, respectively. Thus, the performance metrics in the validation cohort were mainly reduced due to cases with low-confidence or inconclusive classification.

## Discussion

Here, we identified CpGs informative for CNS tumor classification and established a basis for the flexible use of targeted DNA methylation data for this purpose. We developed a machine learning-based strategy to reduce the number of informative features while maintaining performance comparable to other classifiers ^4,32–34^, using only 163-1003 features. The selected features were linked to genes associated with the nervous system and development and may provide opportunities for further biological investigation. Importantly, the approach was compatible with sequencing-based data, increasing its potential for cost-effective CNS tumor classification in clinical practice.

Recent findings suggest that high AUC and low misclassification rates can be achieved using elastic-net methods and linear-kernel SVM classifiers ^35^. SVM-based classification also provided accurate results in our study, with performance maintained after reducing the feature set to 163 features, indicating robustness. Classification was driven by combinations of features spanning several classes rather than individual class-specific features, which may contribute to this robustness. Nevertheless, certain features accurately distinguished selected tumor types.

The potential of DNA methylation sequencing for tumor detection and diagnostics has been demonstrated previously. For example, cfRRBS, also used in our study, enables cost-effective sequencing of the CpG-rich fraction of the genome from low sample inputs, supporting potential clinical applications ^23,36^. Protocols have also been optimized for lower DNA input requirements and plasma cfDNA ^37,38,39,40^. Together, our findings support the feasibility of DNA methylation-based CNS tumor classification using sequencing approaches compatible with more widely available instrumentation. EM-seq and Nanopore sequencing offer cost-efficient, low-input-compatible alternatives to Illumina microarrays with comparable hands-on labour per sample. However, target enrichment being the dominant cost of EM-seq, the total cost of the method depends on the number of samples pooled, creating a trade-off between per-sample cost and turnaround time. Nanopore sequencing currently enables intraoperative classification, whereas short-read approaches do not; for example, we estimate EM-seq to have an estimated minimum turnaround time of six days from sample to result ^41,42^. Faster turnaround with Nanopore, however, may come at a higher per-sample cost ^43^.

A recent study ^34^ introduced a neural network-based CNS tumor classifier using binarized beta values and random masking to simulate sparse methylation profiles. While our results were comparable, our approach uses a hybrid feature-selection strategy integrating consistency filtering with ENLR, enabling identification of compact and biologically meaningful CpG regions. Our classifier maintained high performance in both microarray and sequencing data even when the feature set was reduced to 163 CpGs. Furthermore, retaining continuous beta values rather than binarizing them preserves the resolution of methylation signals, which may be particularly relevant in low-purity tumors where threshold-based binarization could obscure subtle but diagnostically informative patterns.

Our SVM framework provides the predicted tumor class together with confidence scores and probability estimates for the three top-ranked tumor types. This output can help distinguish decisive classifications from potentially ambiguous cases, including those with low tumor content or mixed or atypical tumor types.

An interpretable approach may be particularly useful when methylation profiling results have low confidence or are inconsistent with histopathological diagnoses. Because our approach uses only hundreds of features, methylation profiles can be readily explored, as demonstrated with the DNT sample (S13). SVM class-wise feature weights may further facilitate such evaluation. As more methylation sequencing data become available, exceptions in methylation profiles associated with genetic alterations, tumor location, patient age, immune infiltration, or necrosis may become increasingly identifiable. The characterization of diagnostically informative features may also support development of targeted assays, such as droplet digital PCR or quantitative PCR, for selected tumor entities. We demonstrated this concept by distinguishing MB WNT tumors from other embryonal tumors and astroblastoma, MN1-altered tumors from embryonal tumors using only two features. Such approaches are more feasible with rather homogeneous tumors with consistent differentially methylated sites, whereas heterogeneous entities such as GBM are likely to be more challenging.

A limitation of this study is the moderate size and class coverage of the sequencing cohorts, particularly for underrepresented tumor entities. However, sequencing data were primarily used for independent validation of sequencing compatibility, while a larger external microarray-derived cohort strengthened performance evaluation. Data imputation provided an opportunity to extend the approach to low-coverage Nanopore sequencing and reduce the need for new models in the presence of missing values. Nevertheless, imputed classifications should be interpreted cautiously when confidence scores are suboptimal, sequencing coverage is compromised, or many values are missing. Additional evidence, including individual informative features, other molecular alterations, and histopathological evaluation, should be considered. Imputation may also artificially increase confidence by making methylation profiles more similar to reference profiles. Therefore, despite retained performance after imputation, covering as many of the 1003 features as possible remains preferable.

We updated the model using the most recent v12.8 class structure and the original training data, enabling comparison with both models. Class-level annotations were used as the training data was insufficient for subclass-level classification. Expanding the training cohort, particularly for novel and underrepresented entities, will be important for comprehensive clinical application. Although our results demonstrate applicability across three bisulfite-equivalent targeted short-read sequencing workflows and Nanopore sequencing, compatibility with other sequencing data types or conversion chemistries, including PacBio, TAPS, and Illumina 5-base approaches, remains to be evaluated.

Taken together, our study demonstrates that traditional machine learning approaches can provide an accurate and interpretable framework for DNA methylation-based tumor classification. By combining hybrid feature selection with SVM classification, we identified compact, biologically meaningful feature sets that maintain high diagnostic accuracy while reducing complexity and cost. The framework remains robust with incomplete CpG coverage and provides probability scores, confidence values, and ranked predictions that may support diagnostic interpretation. In addition to classification, EM-seq provides information on diagnostically informative DNA copy number and *MGMT* promoter methylation patterns, potentially supporting treatment stratification. Overall, this work provides a scalable and clinically adaptable framework for precision neuro-oncology beyond current SoC, with potential advantages in cost, TAT, robustness, and interpretability. It uncovers the simplicity and interpretability of DNA methylation-based CNS tumor diagnostics.

## Data Availability

The data supporting the findings of this study are available in the Gene Expression Omnibus (GEO) repository under accession number GSE262533. The code used for analysis is available on GitHub at https://github.com/CRI-group/DNA_meth_brain_tumors. Further inquiries regarding the data and code can be directed to the corresponding author.

## Required statements

## Ethics

The study was performed in accordance with the principles of the Declaration of Helsinki and the guidelines of the Finnish National Board on Research Integrity. This study was approved by the Ethical Committee of Tampere University Hospital (TAYS) (decision R07042 for the retrospective study, dates 9.2017 and 12.2024, and decisions R14024 and R13050 for the adult and pediatric prospective studies, respectively) and the National Supervisory Authority for Welfare and Health (Valvira, decision V/78697/2017). For cases that were part of the retrospective study, the requirement for informed consent was waived by The Regional Ethics Committee of Tampere University Hospital due to the retrospective nature of the study.

## Authorship

Conceptualization: E.M., S.A., K.N., H.H., J.H., K.J.R.; Methodology: E.M., J.V., T.H., A.R.-M., A.D.K., R.V.P., B.D.W., N.C., K.J.R.; Software: E.M., T.H., A.H. F.T.; Validation: E.M.; Formal analysis: E.M., T.H., A.H., M.P., V.F., F.T.; Investigation: J.V., T.H., A.H., L.H., M.P., M.V., A.R.-M., K.J.R.; Resources: S.A., A.M., M.E.L.K., H.-R.T., O.K., K.N., H.H., J.H., M.N., K.J.R.; Data curation: T.H., A.H., M.P., M.V., S.L., K.J.R.; Writing – original draft: E.M., J.V., A.R.-M., K.J.R.; Writing – review & editing: E.M., J.V., T.H., A.H., L.H., A.R.-M., M.P., M.V., S.L., V.F., F.T., S.A., A.M., A.D.K., R.V.P., B.D.W., N.C., M.E.L.K., H.-R.T., O.K., K.N., H.H., J.H., M.N., J.K., K.J.R.; Visualization: E.M., J.V., T.H., A.R.-M., M.P., A.H., K.J.R.; Supervision: A.R.-M., M.P., A.D.K., R.V.P., B.D.W., N.C., M.N., J.K., K.J.R.; Project administration: K.J.R.; Funding acquisition: M.N., K.J.R.

## Acknowledgements

We would like to acknowledge Ms. Laura Nevaharju-Sarantis, Ms. Paula Kosonen, Mrs. Päivi Martikainen, Mrs. Marja Pirinen, Mrs. Sari Toivola, and Mrs. Hanna Selin for sample handling, logistics and skillful technical assistance. We thank Ms. Serafiina Ohlsbom and other key project members in ^44^ for sharing the reference DNA copy number profile plots for EM-seq vs. whole-genome sequencing comparison analysis as well as Dr. Martin Sill and other corresponding authors of ^11^ for sharing the sample-wise MNP v12.8 class labels and confidence scores related to the samples in the original Capper et al. cohort ^4^. Anne Simi and other personnel at Tampere University Hospital and Fimlab laboratories Ltd. are acknowledged for their contribution to sample collection and handling. We are grateful to them and the patients for permitting the analysis of precious patient material. We acknowledge Mrs. Katleen De Preter for her input related to cfRRBS-based analysis. Computational analyses were mostly run on the servers provided by Tampere Center for Scientific Computing, Tampere University, and Bioinformatics Center (https://bioinformatics.uef.fi/biowhat/BIC/bic_101/), University of Eastern Finland, Finland. We acknowledge the CSC - IT Centre for Science, Finland, for providing computational resources as well as the Biocenter Finland (BF) and Tampere Genomics Facility for the service. During the preparation of this manuscript, the authors used ChatGPT (OpenAI) in order to proofread and shorten part of the text. After using this tool, the authors reviewed and edited the content as needed and take full responsibility for the content of the published article.

## References

1. Louis DN, Perry A, Wesseling P, et al. The 2021 WHO Classification of Tumors of the Central Nervous System: a summary. Neuro Oncol. 2021;23(8):1231–1251. doi:10.1093/neuonc/noab106

2. van den Bent MJ. Interobserver variation of the histopathological diagnosis in clinical trials on glioma: a clinician’s perspective. Acta Neuropathol. 2010;120(3):297–304. doi:10.1007/s00401-010-0725-7

3. Leo P, Lee G, Shih NNC, Elliott R, Feldman MD, Madabhushi A. Evaluating stability of histomorphometric features across scanner and staining variations: prostate cancer diagnosis from whole slide images. J Med Imaging (Bellingham). 2016;3(4):047502. doi:10.1117/1.JMI.3.4.047502

4. Capper D, Jones DTW, Sill M, et al. DNA methylation-based classification of central nervous system tumours. Nature. 2018;555(7697):469–474. doi:10.1038/nature26000

5. Louis DN, Perry A, Reifenberger G, et al. The 2016 World Health Organization Classification of Tumors of the Central Nervous System: a summary. Acta Neuropathol. 2016;131(6):803–820. doi:10.1007/s00401-016-1545-1

6. Pajtler KW, Witt H, Sill M, et al. Molecular Classification of Ependymal Tumors across All CNS Compartments, Histopathological Grades, and Age Groups. Cancer Cell. 2015;27(5):728–743. doi:10.1016/j.ccell.2015.04.002

7. Hovestadt V, Remke M, Kool M, et al. Robust molecular subgrouping and copy-number profiling of medulloblastoma from small amounts of archival tumour material using high-density DNA methylation arrays. Acta Neuropathol. 2013;125(6):913–916. doi:10.1007/s00401-013-1126-5

8. Sahm F, Schrimpf D, Stichel D, et al. DNA methylation-based classification and grading system for meningioma: a multicentre, retrospective analysis. Lancet Oncol. 2017;18(5):682–694. doi:10.1016/S1470-2045(17)30155-9

9. Jaunmuktane Z, Capper D, Jones DTW, et al. Methylation array profiling of adult brain tumours: diagnostic outcomes in a large, single centre. Acta Neuropathol Commun. 2019;7(1):24. doi:10.1186/s40478-019-0668-8

10. Priesterbach-Ackley LP, Boldt HB, Petersen JK, et al. Brain tumour diagnostics using a DNA methylation-based classifier as a diagnostic support tool. Neuropathol Appl Neurobiol. 2020;46(5):478–492. doi:10.1111/nan.12610

11. Sill M, Schrimpf D, Patel A, et al. Advancing CNS tumor diagnostics with expanded DNA methylation-based classification. Cancer Cell. 2026;44(2):340–354.e2. doi:10.1016/j.ccell.2025.11.002

12. Taylor AM, Lombardi JT, Patel A, et al. A feasibility study of enzymatic methylation sequencing of cell-free DNA from cerebrospinal fluid of pediatric central nervous system tumor patients for molecular classification. Neurooncol Adv. 2025;7(1):vdaf159. doi:10.1093/noajnl/vdaf159

13. Smith KS, Fischer TT, Han K, et al. M-PACT leverages cell-free DNA methylomes to achieve robust classification of pediatric brain tumors. Nat Cancer. 2026;7(4):667–683. doi:10.1038/s43018-026-01115-4

14. Aryee MJ, Jaffe AE, Corrada-Bravo H, et al. Minfi: a flexible and comprehensive Bioconductor package for the analysis of Infinium DNA methylation microarrays. Bioinformatics. 2014;30(10):1363–1369. doi:10.1093/bioinformatics/btu049

15. Cortes C, Vapnik V. Support-vector networks. Mach Learn. 1995;20(3):273–297. doi:10.1007/BF00994018

16. Boser BE, Guyon IM, Vapnik VN. A training algorithm for optimal margin classifiers. In: Proceedings of the Fifth Annual Workshop on Computational Learning Theory. COLT ’92. Association for Computing Machinery; 1992:144–152. doi:10.1145/130385.130401

17. Vapnik VN. Statistical Learning Theory. Wiley; 1998.

18. Krijthe JH. Rtsne: T-Distributed Stochastic Neighbor Embedding Using Barnes-Hut Implementation.; 2015. https://github.com/jkrijthe/Rtsne

19. Capper D, Stichel D, Sahm F, et al. Practical implementation of DNA methylation and copy-number-based CNS tumor diagnostics: the Heidelberg experience. Acta Neuropathol. 2018;136(2):181–210. doi:10.1007/s00401-018-1879-y

20. Neftel C, Laffy J, Filbin MG, et al. An Integrative Model of Cellular States, Plasticity, and Genetics for Glioblastoma. Cell. 2019;178(4):835–849.e21. doi:10.1016/j.cell.2019.06.024

21. WHO Classification of Tumours Editorial Board. Central Nervous System Tumours: WHO Classification of Tumours Series, 5th Edition. Vol 6. International Agency for Research on Cancer; 2021. https://publications.iarc.fr/601

22. Stone TJ, Merve A, Valerio F, Yasin SA, Jacques TS. Paediatric low-grade glioma: the role of classical pathology in integrated diagnostic practice. Childs Nerv Syst. 2024;40(10):3189–3207. doi:10.1007/s00381-024-06591-6

23. Koker AD, Van Paemel R, Wilde BD, Preter KD, Callewaert N. A versatile method for circulating cell-free DNA methylome profiling by reduced representation bisulfite sequencing. bioRxiv. Published online June 11, 2019:663195. doi:10.1101/663195

24. Bady P, Sciuscio D, Diserens AC, et al. MGMT methylation analysis of glioblastoma on the Infinium methylation BeadChip identifies two distinct CpG regions associated with gene silencing and outcome, yielding a prediction model for comparisons across datasets, tumor grades, and CIMP-status. Acta Neuropathol. 2012;124(4):547–560. doi:10.1007/s00401-012-1016-2

25. Malley DS, Hamoudi RA, Kocialkowski S, Pearson DM, Collins VP, Ichimura K. A distinct region of the MGMT CpG island critical for transcriptional regulation is preferentially methylated in glioblastoma cells and xenografts. Acta Neuropathol. 2011;121(5):651–661. doi:10.1007/s00401-011-0803-5

26. Louis DN, Perry A, Wesseling P, et al. The 2021 WHO classification of tumors of the central nervous system: A summary. Neuro Oncol. 2021;23(8):1231–1251. doi:10.1093/neuonc/noab106

27. Perez G, Barber GP, Benet-Pages A, et al. The UCSC Genome Browser database: 2025 update. Nucleic Acids Res. 2025;53(D1):D1243–D1249. doi:10.1093/nar/gkae974

28. Wei S, Dai M, Liu Z, et al. The guanine nucleotide exchange factor Net1 facilitates the specification of dorsal cell fates in zebrafish embryos by promoting maternal β-catenin activation. Cell Res. 2017;27(2):202–225. doi:10.1038/cr.2016.141

29. Li W, Wang D, Sun X, Zhang Y, Wang L, Suo J. ADAM17 promotes lymph node metastasis in gastric cancer via activation of the Notch and Wnt signaling pathways. Int J Mol Med. 2019;43(2):914–926. doi:10.3892/ijmm.2018.4028

30. Zheng SL, Li ZY, Song J, Liu JM, Miao CY. Metrnl: a secreted protein with new emerging functions. Acta Pharmacol Sin. 2016;37(5):571–579. doi:10.1038/aps.2016.9

31. Alkhanjari RR, Alhajeri MM, Bhamidimarri PM, et al. Septins in the nervous system: from cytoskeletal dynamics to neurological disorders. Cell Commun Signal. 2025;23(1):425. doi:10.1186/s12964-025-02430-6

32. Chen Y, Yan Y, Xu M, et al. Development of a Machine Learning Classifier for Brain Tumors Diagnosis Based on DNA Methylation Profile. Front Bioinform. 2021;1:744345. doi:10.3389/fbinf.2021.744345

33. Santana-Santos L, Kam KL, Dittmann D, et al. Validation of Whole Genome Methylation Profiling Classifier for Central Nervous System Tumors. J Mol Diagn. 2022;24(8):924–934. doi:10.1016/j.jmoldx.2022.04.009

34. Yuan D, Jugas R, Pokorna P, et al. crossNN is an explainable framework for cross-platform DNA methylation-based classification of tumors. Nat Cancer. 2025;6(7):1283–1294. doi:10.1038/s43018-025-00976-5

35. Maros ME, Capper D, Jones DTW, et al. Machine learning workflows to estimate class probabilities for precision cancer diagnostics on DNA methylation microarray data. Nat Protoc. 2020;15(2):479–512. doi:10.1038/s41596-019-0251-6

36. Van Paemel R, De Koker A, Vandeputte C, et al. Minimally invasive classification of paediatric solid tumours using reduced representation bisulphite sequencing of cell-free DNA: a proof-of-principle study. Epigenetics. 2021;16(2):196–208. doi:10.1080/15592294.2020.1790950

37. Shen SY, Burgener JM, Bratman SV, De Carvalho DD. Preparation of cfMeDIP-seq libraries for methylome profiling of plasma cell-free DNA. Nat Protoc. 2019;14(10):2749–2780. doi:10.1038/s41596-019-0202-2

38. Taiwo O, Wilson GA, Morris T, et al. Methylome analysis using MeDIP-seq with low DNA concentrations. Nat Protoc. 2012;7(4):617–636. doi:10.1038/nprot.2012.012

39. Lak NSM, van Zogchel LMJ, Zappeij-Kannegieter L, et al. Cell-Free DNA as a Diagnostic and Prognostic Biomarker in Pediatric Rhabdomyosarcoma. JCO Precis Oncol. 2023;7:e2200113. doi:10.1200/PO.22.00113

40. Nassiri F, Chakravarthy A, Feng S, et al. Detection and discrimination of intracranial tumors using plasma cell-free DNA methylomes. Nat Med. 2020;26(7):1044–1047. doi:10.1038/s41591-020-0932-2

41. Djirackor L, Halldorsson S, Niehusmann P, et al. Intraoperative DNA methylation classification of brain tumors impacts neurosurgical strategy. Neurooncol Adv. 2021;3(1):vdab149. doi:10.1093/noajnl/vdab149

42. Vermeulen C, Pagès-Gallego M, Kester L, et al. Ultra-fast deep-learned CNS tumour classification during surgery. Nature. Published online October 11, 2023. doi:10.1038/s41586-023-06615-2

43. Halldorsson S, Nagymihaly RM, Bope CD, et al. Whole-genome long-read sequencing for rapid comprehensive molecular diagnostics of brain tumors. medRxiv. Published online April 24, 2026:2026.04.23.26351563. doi:10.64898/2026.04.23.26351563

44. Ohlsbom S, Mäntylä S, Nätkin R, et al. Multi region whole genome and transcriptomic profiling uncovers plastic, subclone linked cell states in high grade diffuse astrocytomas. bioRxiv. Published online August 11, 2026:2026.08.11.743949. doi:10.64898/2026.08.11.743949

