## Supplementary Methods for "Consistent DNA methylation patterns enable accurate and interpretable cross-platform classification of central nervous system tumors"

**Preprocessing and normalization of the microarray data**

We filtered, batch-effect corrected, and normalized the data as described by Capper et al. ^1^, and calculated detection p-values. The Capper method included the following probe filters for removal of: 1) probes targeting sex chromosomes; 2) probes containing a single-nucleotide polymorphism (dbSNP132 Common) within five base pairs of and including the targeted CpG site; 3) probes not uniquely mapped to the human reference genome (hg19), allowing for one mismatch; and 4) probes not included in the Illumina EPIC array. No samples were dropped based on p-values of <0.05. We refrained using the detection p-value for probe filtering.

For building a validation cohort, we similarly downloaded and pre-processed the validation dataset of Capper et al. (GSE109379) from GEO, which resulted in an additional 1104 samples and 428,799 probes. We then constructed another validation set from six cohorts (GSE147548, GSE125450, GSE198855, GSE156090, GSE193196, and GSE104210), resulting in 899 samples and 428,230 probes. GSE337222 data set was processed similarly for validating the binary classification results for separating MB WNT from other embryonal tumors. We determined methylation-based classes for samples without prior classification results available using the next-generation molecular neuropathology (MNP) v11b4 (version 3.3) platform that was hosted by the German Cancer Research Center (DKFZ) and later, for comparison purposes, version v12.8 accessible via Epignostic (<https://app.epignostix.com/>). The v12.8 class labels and MNP confidence scores of samples within the Capper et al training and validation set were kindly provided by the corresponding author of the original publication, Dr. Martin Sill ^2^.

### **Consistency filtering with intra- and multi-class approaches**

A hybrid feature selection strategy, which consists of a combination of several filtering approaches and a supervised feature selection technique known as Elastic Net Logistic Regression (ENLR), was constructed for refining the original feature set (see Supplementary Figure S1A). The goal was to simultaneously improve the classification performance and refine the feature set to include only the most relevant features. The intra-class consistency and multi-class consistency approaches were selected in order to balance between intra-class consistency and proportion of inconsistent classes. The intra class consistency approach required a higher intra-class consistency, yet allowing a higher proportion of inconsistent classes, whereas multi-class consistency approach allowed lower intra-class consistency but required higher numbers of consistent classes instead (Supplementary Figure S1A).

First, the 32,000 most variable probes were selected from the Capper et al. ^1^ reference set in combination with blood and cfDNA sample sets. Blood and cfDNA sets were excluded from further analysis. We first determined the β-value group (group 1: beta≤0.3, group 2: 0.3<beta<0.7, and group 3: beta≥0.7) for each selected feature across samples. We then further categorized tumor classes to consistently unmethylated (“1”), consistently intermediately methylated (”2”), consistently methylated (”3”) or non-consistently methylated (”0”) based on the features methylation status consistency. Intra-class consistency approach required the features determined group (“1”,”2”,”3”) to be present consistently in ≥90% of the samples of that tumor class to determine feature as consistent, whereas 80% was used as a consistency requirement in the multi-class consistency approach. The features failing to reach these limits were assigned as inconsistent (“0”). We next further filtered the feature set with the following inclusion criteria for the consistent features: both consistent methylation status extremes “1” and “3” were required to be present at a retained feature across tumor classes or at least two categories to be present in a retained feature across classes for the intra- and multi-class consistency approaches respectively. As a final filtering step we removed the features containing non-consistent category classes in >55% (n≥50) classes and in >30% (n≥27) classes respectively. Filtering resulted in 4,109 features and 1,826 features for intra-class consistency approach and multi-class consistency approach, respectively.

### **ENLR-based feature selection**

The initial set of 450,000 features was reduced to 4,976 following filtering-based feature selection. To further refine the feature selection process, we employed ENLR ^3^ that combines LASSO and ridge penalties ^4^. We implemented ENLR using the glmnet package (version 4.1-7) in R (version 4.1.1). To briefly explain ENLR, the LASSO penalty acts as a feature selector by enforcing many parameters to have zero values, thus leading to a sparse solution. In applications with highly correlated features, LASSO tends to select only one of them while ignoring the other correlated features, even if they are relevant. By contrast, a ridge regression penalty shrinks the coefficients of the correlated features toward each other and assigns similar coefficient values to them. However, ridge regression does not yield sparse solutions. A combination of these two penalties, that is, the elastic net penalty, leads to a sparse model combined with the grouping effect. This renders the approach particularly suitable for applications involving highly correlated features.

ENLR is a supervised approach that incorporates label information during the feature selection process. We applied ENLR using alpha value 0.5 within the cross-validation loop before implementing our actual classification approach (SVM) (Figure 1C). The ENLR feature selection step was performed using only the training data in each cross-validation split. However, the initial variance- and consistency-based filtering steps were performed once on the complete training cohort before cross-validation. Consequently, although ENLR was applied within the cross-validation loop, the internal cross-validation results should be interpreted as internal performance estimates rather than unbiased estimates of generalization performance. Therefore, the independent external validation cohort was used as the primary assessment of classifier generalizability.

For the ENLR-based feature selection, we conducted 10 runs of 10-fold cross-validation, resulting in 100 models. We selected features based on an average coefficient value across the 100 models as well as the top most important features per class. To further assess the stability of the selected 1003-feature panel across repeated ENLR runs, we quantified feature overlap between models using Jaccard similarity indices and evaluated coefficient variability across the 100 ENLR models.

To evaluate the performance of our hybrid feature selection approach, we generated 10 random feature panels, each containing 1,003 high-variance probes matching the size of the selected feature panel. Using the same training and external validation pipeline, the selected 1,003 features achieved an external validation accuracy of 88.8% (using MNP class labels as ground truth), whereas the random feature panels achieved a mean accuracy of 46.9% (range, 46.3%–47.5%). These results demonstrate that our hybrid feature selection approach identifies substantially more informative features than a variance-matched random feature set of the same size.

### **Annotation and Gene set enrichment analysis (GSEA)**

We annotated the features in the 1003 set and the 4976 set to the nearest gene according to the location with respect to the gene, using the pre-built hg19 annotations of the annotatr Bioconductor package ^5^ with default settings. We then performed a gene set enrichment analysis (GSEA) separately for genes having probe(s) at the promoter, other genomic region(s) or either (group “All”), as in ^6^. Terms with an enrichment p-value greater than 0.001, fold enrichment below two, and containing fewer than 10 genes were filtered out. The group “All'' comprises all the genes that have a selected feature in any of the following locations: proximal promoter (0-1,000 bp upstream from the TSS), introns, exons, 5’UTR, 3’UTR and distal promoter (1,000-5,000 bp upstream from the TSS). The category “All -Promoters'' includes those genes with a measured feature in any of the above locations except the proximal and distal promoters. The category “Promoters” consists of those genes that have a feature in the proximal and/or the distal promoters. We reduced the redundancy from the term lists with the help of Revigo ^7^. Lastly, we categorized the biological process Gene Ontology (GO) terms manually into themes.

### **Classifier construction**

We used a multiclass SVM with a linear kernel to build the prediction model. Multiclass SVM is based on pairwise SVM classifications, contrasting each DNA methylation class with all other classes. The final classification decision is based on the majority vote (i.e. which tumor class is selected most frequently). In more detail, SVMs were first introduced ^8–10^ as a pattern recognition method representing decision boundaries between samples from two different classes, such that the margin (distance) between the decision boundary and the closest training sample to it is maximized. The SVM transforms the training data from the original space into a high-dimensional feature space using a kernel-induced mapping function, and the separating hyperplane is computed in this new feature space. Although the SVM classification approach is primarily designed for binary classification, it can be utilized for multiclass classification problems by breaking them down the multi-classification problem into multiple binary classification problems. Our select one-on-one approach treats the multiclass problem as a multiple binary classification problem with one binary classifier for each pair of classes. The decision values were used to predict the class labels.

A set of 10 samples from Capper’s validation cohort yielded a misleading profile according to the original publication ^1^, and were not included in the validation dataset. This indicates that these samples showed a high MNP classifier confidence score and a conflicting methylation profile compared to the original tumor diagnosis, yet tumor classification was not altered after pathological re-evaluation. Of note, SVM did not classify these samples any better than the MNP classifier.

### **Pathological evaluation of the in house patient cohort**

An experienced neuropathologist evaluated the FFPE tumor samples and determined the histopathological type and grade according to the WHO criteria valid at diagnosis. In addition, tumor location, history, and ancillary studies (e.g., immunohistochemistry, next-generation sequencing, PCR) were used for tumor diagnosis whenever available. Frozen tumor sections were stained with hematoxylin and eosin (H&E) to estimate the tumor content in the samples before DNA isolation.

### **Preparation of sequencing libraries and sequencing data analysis**

The EM-seq libraries were prepared according to the EM-seq workflow ^11^ using a reagent kit and a protocol for use with standard insert libraries from NEB, with few deviations. Briefly, 110 - 140 ng of each sample was sheared to approximately 200 bp using a Covaris S220 instrument to obtain a target bp (peak) of 150 with using following settings: Peak incident power 175W; Duty factor 10%; Cycles per Burst 200; Treatment time: 450s (Covaris, Massachusetts, USA). The fragmentation profiles were monitored using a High-Sensitivity D1000 ScreenTape on a 4150 TapeStation Instrument (Agilent Technologies) and concentration was measured with Qubit dsDNA High Sensitivity assay or Qubit 4 fluorometer (Invitrogen, Thermo Fisher Scientific). A total of 45 to 148 ng of each sample was further subjected to the EM-seq workflow. In between deamination of cytosines and clean-up of deaminated DNA, the samples were stored overnight at 4°C. The rest of the protocol was carried out as instructed, using eight to ten cycles of PCR. Quality control of the fragmentation profile of the libraries was performed using a High Sensitivity D1000 ScreenTape on a 4150 TapeStation Instrument (Agilent Technologies), in addition to concentration measurements with a Qubit HS dsDNA assay and Qubit 4 fluorometer (Invitrogen, Thermo Fisher Scientific). Target enrichment was performed with a IDTs product “custom xGen Custom Hyb Panel-Accel” covering a total of 1124 targets: 1003 CpGs and 121 other selected areas, such as MGMT promoter region and *CDKN2A* locus. Hybridization was carried out using the IDT xGen Hybridization Capture of DNA Libraries Protocol and Appendix A of the protocol with v2 reagents for the batch 1 samples (S1-S20) and with v3 for batch 2 samples (S23-S57). Briefly, the target enrichment was carried out with 5 to 12 libraries per pool with varying inputs up to 500 ng per library and using a 4h hybridization time followed by 10-14 cycles of post-capture PCR. Library quality was estimated using the Qubit dsDNA Broad Range assay and Qubit 4.0 instrument (Invitrogen) and TapeStation instrument with ScreenTape D1000 (Agilent Technologies). Samples S1-S20 were sequenced with Illumina Novaseq 6000 using external sequencing provider to an average 6.5 M [range 4.6-8.0 M] reads per sample. Samples S23-S57 were sequenced in-house with AVITI (Element Bio) using AVITI 2x150 Sequencing Kit Cloudbreak FS Low Output (860-00011) to 8.5 M [range 2.1-20.1 M] reads per sample.

A custom capture library containing 46,084 probes (2.923 Mbp) was designed using the SureSelect DNA Advanced Design Wizard (Agilent Technologies) on the GRCh37/hg19 genome build. The SureSelect libraries were prepared using the SureSelectXT Methyl reagent kit (Agilent Technologies) according to the manufacturer’s protocol for 1 µg DNA samples (SureSelectXT Methyl-Seq Target Enrichment System for Illumina Multiplexed Sequencing), with few exceptions. Briefly, 600 ng of input DNA per sample was sheared to approximately 170 bp, using a Covaris S220 instrument (Covaris, Massachusetts, USA). Hybridization was performed for 16 h according to the above mentioned manufacturer’s protocol, with the following exception. Due to reduced input DNA, only 2 µl of capture library were used per reaction, with 5 µl of 10% RNase BLock solution. Bisulfite conversion was performed using a Premium Bisulfite kit (Diagenode, Liège, Belgium) according to the manufacturer’s instructions. Bisulfite-treated libraries were amplified using 13 cycles, and Indexing PCR was performed as indicated in the Agilent protocol.

cfRRBS libraries were done using the original protocol ^12,13^ with 10 ng of sample DNA input appropriately mixed with unmethylated lambda DNA controls but deviating from the original protocol by using Diagenode’s Premium Bisulfite Conversion Kit (Diagenode, Liège, Belgium) for the bisulfite conversion step. The resulting Sureselect and cfRRBS libraries were sequenced on an Illumina NovaSeq sequencer (Illumina) using a PE150 option.

Performing target enrichment after library preparation enables utilization of the full capabilities of novel sequencers, such as the AVITI’s ability to perform the enrichment on the flow cell, but it also reduces the DNA input requirement compared to the requirements of array hybridization or SureSelect-based sequencing and a small targeted panel requires less sequencing than other less focused sequencing methods such as RRBS ^14^ or MeDIP ^15^.

For EM-seq, SureSelect Methyl-seq, and cfRRBS libraries, library quantity was monitored with Qubit dsDNA Broad Range assay or High Sensitivity assay using Qubit™ 4.0 instrument (Invitrogen). Library quality was monitored with TapeStation instrument and D1000 or High-Sensitivity D1000 Screen tape (Agilent Technologies) or alternatively with Fragment Analyzer 5200 and HS NGS Kit (Agilent Technologies).

We processed the EM-seq and the SureSelect data files similarly. First, we conducted a sequencing data QC step with FastQC, trimmed adapter sequences with Trim Galore, performed bisulfite mapping against hg19 (UCSC), and conducted a mapping quality control with duplicate removal using Bismark v0.22.3 ([https://www.bioinformatics.babraham.ac.uk](https://www.bioinformatics.babraham.ac.uk/)) ^16^. Methylation extraction was performed in paired-end mode using the --comprehensive setting. M-bias profiles were inspected, and the first two bases of Read 2 were excluded from methylation calling (--ignore_r2 2). For processing the cfRRBS data, we used an in-house Snake-make pipeline – MethylSnake ([https://github.com/ftabaro/MethylSnake](https://github.com/ftabaro/MethylSnake/blob/master/singularity/Singularity.base)) applying the same tools and steps as those used for EM-seq and SureSelect data with the exception that the duplicate removal phase was omitted since it is not applicable to RRBS data without UMI information.

MethylKit 1.16.1 ^17^ was used in R 4.0.4 environment (R Core Team 2021) for coverage filtering, removing CpGs with read coverage <10, and merging reads on both strands of a CpG dinucleotide when uniting samples. Read coverage of 10 was selected to control the noise level of beta value calculation without losing too many CpGs. To represent the methylation level for each probe, we used the probe coordinates from Illumina HumanMethylation 450 k array (2.0.6) to select the measured CpGs. For the additional EMSeq samples (S23-S57), a similar workflow was implemented using Bismark v. 0.24.2, R version 4.4.1 and methylKit version 1.32.1. For the SVM models for which we had less than 1003 features and we omit using imputation, the methylation level for each feature was calculated by using the probe coordinates from Illumina HumanMethylation 450 k array (2.0.6) to select the nearest measured CpG within 200 bp. We then expanded the coordinates by 50 bp in both directions and calculated the mean methylation level of CpGs within this 100bp region as an input to the classifier (Supplementary Table S3).

Beta values for index CpGs with insufficient coverage were estimated using K-nearest neighbors imputation. Missing values were replaced with the median beta value of seven most similar samples, identified with Pearson correlation, from combined reference and validation datasets (GSE109381) of Capper et al. (2018).

### **Nanopore data preprocessing**

Methylation data, including 259 measurements from 175 CNS tumors, was obtained from the publicly available dataset deposited at Zenodo (https://doi.org/10.5281/zenodo.13236096), which contain methylation values and measurement probabilities from Nanopore sequencing datasets deposited to European Genome-Phenome Archive (accession no. EGAD50000000832 and EGAD50000000791).

The methylation values matching 1003 CpGs were retained and the replicate samples within the two data sets were combined to increase feature coverage and depth, resulting in 208 measurements. The mean methylation probability of CpG sites was used as a surrogate for beta values, accounting for uncertainty in methylation calls and providing more similar beta values than discrete 0/0.5/1 calls for most measurements, as the sequencing coverage for 44.6% of CpGs was 1 or 2 reads.

**Copy number alterations from targeted sequencing data**

Targeted sequencing generates low-coverage genome-wide data through off-target capture and imperfect isolation of target sequences. Reads within 500bp of target probe sequences were excluded and sample CNAs estimated from the preprocessed and aligned EM-seq data with R-package QDNAseq (v1.42.0) using 500kb binning (<https://pmc.ncbi.nlm.nih.gov/articles/PMC4248318/>). The standard QDNAseq pipeline was followed, including exclusion of problematic regions, correction for GC and mappability biases, bin normalization, removal of outlier bins, and segmentation of chromosomal bins using a square-root transformation. The segmented copy number calls were adjusted for tumor purity, (copy number-2*(1-purity)) / purity, using tumor fraction estimates determined by a neuropathologist from the respective H&E-stained sample tissues.

To infer CNAs in regions targeted for EM-seq enrichment, read counts overlapping with the 500bp extended targeted sites were extracted and total count normalized. Tumor copy numbers were estimated from the ratio of read depths at query sites of tumor samples against the median read depth at the query site of WBC control samples of the batch (n=3). Observed copy numbers were adjusted for tumor purity similarly to the above.

**Clinically meaningful methylation class separation with few consistent features**

Leveraging the selected 1003 feature set, we sought to find features that are consistent amongst compared groups but show differential diagnostics enabling DNA methylation levels for a diagnostically relevant class within histologically similar tumor classes. Moreover, we focused on finding features that distinguish “Medulloblastoma, WNT-activated” (MB, WNT) from other embryonal tumors and high-grade neuroepithelial tumors with MN1 alterations” (HGNET, MN1, currently diagnosed as astroblastoma, MN1-altered) from embryonal tumors.

We selected features based on their determined consistency criteria (Supplementary Table S2), aiming to find features that are consistent in nature in sample groups that are compared but differentially methylated in the index methylation class vs. the references. Next, we plotted beta values of selected probes across all the included tumor types in the reference cohort. The beta value plots of the reference cohort were used to determine viable beta value thresholds for separating the index tumor group from the rest, and performance metrics of differential diagnostics were calculated using these. The tested cutoffs were either beta ≤ 0.3 and/or beta ≤ 0.5, based on beta value plots’ visual guidance. We obtained better performance when we required both selected probes to fulfil the criteria, and thereby this requirement was used in the final analysis. After determining the thresholds based on the performance metrics in the reference cohort, we validated the feasibility of our selection criteria by repeating the performance metric calculations for the validation cohort and plotted the beta values similarly to the reference cohort.

To further validate our results we processed an external GEO dataset (GSE337222) and presented the results of this analysis similarly to the above (Supplementary Figure S16). Statistical analyses were performed in Python (v3.14.5) using NumPy (v2.4.4) and pandas (v3.0.2). Ninety-five percent confidence intervals for accuracy, precision, and recall were calculated using Wilson score intervals, and confidence intervals for the F1 score were estimated by nonparametric bootstrap resampling (100,000 iterations). Furthermore, we inspected the location of the CpGs that the selected probes measure in respect to genes and the ENCODE annotations with the UCSC Genome Browser (https://genome.ucsc.edu) ^18^.

**tSNE visualization**

We plotted the EM-seq samples together with the reference cohort samples for tSNE visualization ^19^. We combined the EM-seq and capper samples into a single matrix using the identified 1003 features. Next, we calculated the distance matrix with the scaled Pearson correlation (1-cor("pearson")) / 2 ) and processed that with Rtsne, using 50 initial dimensions, perplexity of 30, and 1000 iterations.

**Cost and Turn-around-time estimations**

The legacy technology The SureSelectXT Methyl-Seq Library Preparation (Agilent) is to be discontinued on November 30th, 2026. Thereby, cost or turn-around estimations regarding this are in a sense irrelevant for future clinical application and thereby not reported. The cost (excluding personnel costs) and turn-around time (TAT) of the cfRRBS methodology have been reported to be 180€ and 5 working days^13^. For targeted EM-seq, the reagent-cost including DNA extraction kit, fragmentation materials, EM-seq library preparation, target enrichment with IDT xGen™ Hybridization Capture Core Reagents and sequencing adds up to 110 - 126 € / sample depending on sequencing instrumentation used.

The cost estimates of targeted EM-seq are based on available information about purchasing costs of DNA extraction kits (3.86 - 8.84 €/sample [Fresh Frozen - FFPE]), fragmentation materials (8.04€ / sample [Covaris microTUBEs]), Library preparation materials from the NEB site, to be 42.30 € / sample including library prep reagents and indexes used in EM-seq workflow. Hybridization capture reagents are estimated based on prices available from IDT’s web page for xGen™ Hybridization Capture Core Reagents used in pools of 8-samples, 52 € / sample. Sequencing is estimated with the options to sequence 6.5M per sample, either 1) with other samples in the most cost-efficient platform Novaseq X 25B flow-cells, or 2) to sequence in -house with AVITI sequencing instrument with other samples. This creates a sequencing cost-range 3.4 - 19.7 € / sample depending on the instrument used.

1) Novaseq X - Illumina, 25B flow cell capable for 6,500 M total reads per lane, 1 sample = 0.1% of capacity) - Cost of a single lane on an external sequencing provider is estimated to cost 3,400€

2) AVITI Element Biotechnology, AVITI 2x150 Sequencing Kit Cloudbreak FS Low Output [860-00011], capable for 250 M total reads per flow cell, 1 sample = 2.6% of capacity, cost of a single flow cell is ~758€ (880 USD) .

For estimating turn-around time. Experience from the two prepared batches were used to estimate how a batch of 8 samples can be operated through the protocol and is now presented below.

**_________________________________________________________________________EM-seq library preparation schedule**

DAY 1 - Fragmentation and library prep day 1
Extraction and fragmentation
8:00–10:00 — DNA extraction (30 min hands-on)
10:00–11:00 — QC (30 min hands-on)
10:30–11:15 — Fragmentation, overlapping with QC (45 min hands-on)
11:30–12:00 — Fragmentation QC (5 min hands-on)

Library construction
12:15–12:30 — Reagent pipetting (15 min hands-on)
12:30–13:00 — A-tailing reaction
13:00–13:15 — Ligation reaction pipetting (15 min hands-on)
13:15–13:30 — Ligation reaction
13:30 — AMPure XP cleanup (30 min hands-on)
14:00 — TET reaction pipetting (30 min hands-on)
14:30–15:30 — TET incubation
15:30–16:00 — TET stopping reaction + pipetting (30 min hands-on)
NOTE: Reaction can be left in the thermal cycler at +4°C — Safe stopping point

DAY 2
8:00–8:30 — AMPure XP cleanup (30 min hands-on)
8:30–8:45 — DNA denaturation reaction pipetting (15 min hands-on)
8:45–9:00 — Denaturation
9:00–9:30 — Deamination reaction pipetting (30 min hands-on)
9:30–10:00 — Deamination reaction
10:00–10:30 — PCR reaction pipetting (30 min hands-on)
10:30–11:00 — PCR reaction (depending on number of cycles)
11:00–11:30 — AMPure XP purification (30 min hands-on)
11:30–12:30 — QC (TapeStation and Qubit) (30 min hands-on)
12:30–13:00 — Pooling for hybridization (30 min hands-on)

DAY 3
8:00–16:00 — Hybridizations and washes (~2h hands-on)

DAYS 4–5
Sequencing with PE150 takes roughly 48h

DAY 6
Data-analysis is estimated to take no more than a workday, most likely a maximum of 3 hours.

**References**

1.	Capper D, Jones DTW, Sill M, et al. DNA methylation-based classification of central nervous system tumours. *Nature*. 2018;555(7697):469-474. doi:[10.1038/nature26000](http://dx.doi.org/10.1038/nature26000)

2.	Sill M, Schrimpf D, Patel A, et al. Advancing CNS tumor diagnostics with expanded DNA methylation-based classification. *Cancer Cell*. 2026;44(2):340-354.e2. doi:[10.1016/j.ccell.2025.11.002](http://dx.doi.org/10.1016/j.ccell.2025.11.002)

3.	Friedman J, Hastie T, Tibshirani R. Regularization Paths for Generalized Linear Models via Coordinate Descent. *J Stat Softw*. 2010;33(1):1-22. doi:[10.1109/TPAMI.2005.127](http://dx.doi.org/10.1109/TPAMI.2005.127)

4.	Zou H, Hastie T. Regularization and variable selection via the elastic net. *J R Stat Soc Series B Stat Methodol*. 2005;67(2):301-320. doi:[10.1111/j.1467-9868.2005.00503.x](http://dx.doi.org/10.1111/j.1467-9868.2005.00503.x)

5.	Cavalcante RG, Sartor MA. Annotatr: Genomic regions in context. *Bioinformatics*. 2017;33(15):2381-2383. doi:[10.1093/bioinformatics/btx183](http://dx.doi.org/10.1093/bioinformatics/btx183)

6.	Eden E, Navon R, Steinfeld I, Lipson D, Yakhini Z. GOrilla: a tool for discovery and visualization of enriched GO terms in ranked gene lists. *BMC Bioinformatics*. 2009;10(1):48. doi:[10.1186/1471-2105-10-48](http://dx.doi.org/10.1186/1471-2105-10-48)

7.	Supek F, Bošnjak M, Škunca N, Šmuc T. REVIGO summarizes and visualizes long lists of gene ontology terms. *PLoS One*. 2011;6(7):e21800. doi:[10.1371/journal.pone.0021800](http://dx.doi.org/10.1371/journal.pone.0021800)

8.	Cortes C, Vapnik V. Support-vector networks. *Mach Learn*. 1995;20(3):273-297. doi:[10.1007/BF00994018](http://dx.doi.org/10.1007/BF00994018)

9.	Boser BE, Guyon IM, Vapnik VN. A training algorithm for optimal margin classifiers. In: *Proceedings of the Fifth Annual Workshop on Computational Learning Theory*. COLT ’92. Association for Computing Machinery; 1992:144-152. doi:[10.1145/130385.130401](http://dx.doi.org/10.1145/130385.130401)

10.	Vapnik VN. *Statistical Learning Theory*.; 1998. <https://books.google.com/books/about/Statistical_Learning_Theory.html?hl=&id=RWrlkQEACAAJ>

11.	Vaisvila R, Ponnaluri VKC, Sun Z, et al. Enzymatic methyl sequencing detects DNA methylation at single-base resolution from picograms of DNA. *Genome Res*. 2021;31(7):1280-1289. doi:[10.1101/gr.266551.120](http://dx.doi.org/10.1101/gr.266551.120)

12.	Koker AD, Van Paemel R, Wilde BD, Preter KD, Callewaert N. A versatile method for circulating cell-free DNA methylome profiling by reduced representation bisulfite sequencing. *bioRxiv*. Published online June 11, 2019:663195. doi:[10.1101/663195](http://dx.doi.org/10.1101/663195)

13.	Van Paemel R, De Koker A, Vandeputte C, et al. Minimally invasive classification of paediatric solid tumours using reduced representation bisulphite sequencing of cell-free DNA: a proof-of-principle study. *Epigenetics*. 2021;16(2):196-208. doi:[10.1080/15592294.2020.1790950](http://dx.doi.org/10.1080/15592294.2020.1790950)

14.	Meissner A, Gnirke A, Bell GW, Ramsahoye B, Lander ES, Jaenisch R. Reduced representation bisulfite sequencing for comparative high-resolution DNA methylation analysis. *Nucleic Acids Res*. 2005;33(18):5868-5877. doi:[10.1093/nar/gki901](http://dx.doi.org/10.1093/nar/gki901)

15.	Taiwo O, Wilson GA, Morris T, et al. Methylome analysis using MeDIP-seq with low DNA concentrations. *Nat Protoc*. 2012;7(4):617-636. doi:[10.1038/nprot.2012.012](http://dx.doi.org/10.1038/nprot.2012.012)

16.	Krueger F, Andrews SR. Bismark: a flexible aligner and methylation caller for Bisulfite-Seq applications. *Bioinformatics*. 2011;27(11):1571-1572. doi:[10.1093/bioinformatics/btr167](http://dx.doi.org/10.1093/bioinformatics/btr167)

17.	Akalin A, Kormaksson M, Li S, et al. methylKit: a comprehensive R package for the analysis of genome-wide DNA methylation profiles. *Genome Biol*. 2012;13(10):R87. doi:[10.1186/gb-2012-13-10-r87](http://dx.doi.org/10.1186/gb-2012-13-10-r87)

18.	Perez G, Barber GP, Benet-Pages A, et al. The UCSC Genome Browser database: 2025 update. *Nucleic Acids Res*. 2025;53(D1):D1243-D1249. doi:[10.1093/nar/gkae974](http://dx.doi.org/10.1093/nar/gkae974)

19.	Krijthe J, van der Maaten L. T-Distributed Stochastic Neighbor Embedding using a Barnes-Hut Implementation. Published online 2023. <https://cran.r-project.org/web/packages/Rtsne/Rtsne.pdf>
