## Supplementary Table and Figure Legends for "Consistent DNA methylation patterns enable accurate and interpretable cross-platform classification of central nervous system tumors"

### Supplementary files captions

**Supplementary Figure S1.**

(A) Schematic representation of the filter-based strategies used. A tumor class-specific methylation category of 1 (unmethylated), 2 (intermediately methylated) or 3 (methylated) was given to a feature if at least a determined percentage of samples (90% or 80%) in that specific tumor class represent the same methylation level (beta ≤0.3, 0.3-0.7 or ≥0.7, respectively). The category 0 (Inconsistent) was given when any of the above conditions was not met. Features were then filtered based on criteria of a minimum of two different categories present and a maximum number of inconsistent tumor classes (50 and 27 classes in intra- and multi-class consistency approaches, respectively).

(B) Bar plot showing the proportions of features (y-axis) falling into different types of genomic locations (x-axis) for the 4976 feature set. Absolute feature counts for each genomic location are shown above the corresponding bar.

(C) Karyoplot representing the distribution of 1003 (red bars) and 4976 (blue bars) features throughout the genome. Visualized 4976 features are those selected with filter-based feature selection before elastic net logistic regression-based feature selection.

(D) Karyoplot representing the features measured with the Illumina 450k methylation array.

**Supplementary Figure S2.**

Most of the DNA methylation classes showed a clearly distinct methylation profile when compared to other classes. Very few or none differences in consistent features were detected between DNA methylation classes inside certain tumor types (ATRT, GBM, non-WNT/non-SHH MB) and within IDH-mutant diffuse gliomas. Heatmap showing a pairwise comparison of the number of features that belong to different consistent methylation categories (1-3) for the 1003 features. Differences between category 0 (inconsistent) and consistent categories are not considered in the analysis. Clustering was done using euclidean distance and complete linkage.

**Supplementary Figure S3.**

(A) Heatmap representation of the classification accuracy based on the reference cohort shows good performance across tumor and normal DNA methylation classes. Somewhat poorer performance was detected in part of the glioblastoma multiforme (GBM) classes, and in DLGNT and PIN T PB A. Only two tumor groups, GBM MES and DLGNT, showed an average balanced accuracy below 0.92, yet remained above 0.86. Total number of samples in the class and the percentage of misclassified samples are shown on the right.

(B) Confusion matrix showing the misclassified samples at the DNA methylation class level for reference cohort. Discrepancy is shown both at the level of DNA methylation classes and after grouping the methylation classes into one normal and 59 tumor sample groups based on the 5th edition of the WHO classification of CNS tumors, hereafter known as diagnostic classes The majority of the misclassifications fall within the same diagnostic class, such as GBMs or MB SHH, or between similar tumor types, such as IDH-mutant gliomas. Six control samples were misclassified as low-grade gliomas (LGGs) and two DLGNTs as pilocytic astrocytomas.

(C) Performance of the classifier during the training phase. Box plot showing the values of overall performance metrics: balanced accuracy, F1 score, precision, and recall. The center line shows the median, the first and third quartiles are represented by box limits and whiskers depict 1.5× interquartile range. The overall performance values were all above 0.97.

(D) A confusion matrix illustrating misclassifications at the diagnostic tumor class level in the reference cohort, using MNP classification as the ground truth. The number of misclassified samples decreased from 64 to 17 when evaluated at the diagnostic class level.

(E) Accuracy of the SVM classification in the validation cohort when using MNP v11b classification as the ground truth. In addition to the overall performance in the whole validation cohort, accuracy was also calculated separately for samples in each MNP confidence group which was determined based on the calibrated score of the MNP Classifier. Group 1 includes samples with a score equal or larger than 0.84 (n=1658) (high-confidence group), group 2 samples with a score 0.5-0.84 (n=241), and group 3 samples with a score smaller than 0.5 (n=104). Samples with a misleading profile (n=10) (as reported in the original publication by Capper *et al*.) were analyzed separately.

(F) Accuracy of the SVM classification in the validation cohort when using MNP v11b classification as the ground truth. In addition to the overall performance in the whole validation cohort, accuracy was also calculated separately for samples in each MNP confidence group which was determined based on the calibrated score of the MNP Classifier. Group 1 includes samples with a score equal or larger than 0.90 (n=1589) (high-confidence group), group 2 samples with a 0.5 <= score < 0.90 (n=300), and group 3 samples with a score smaller than 0.5 (n=104). Samples with a misleading profile (n=10) (as reported in the original publication by Capper *et al*.) were analyzed separately.

(G) Performance metrics in the validation cohort for each DNA methylation class for samples belonging to MNP group 1.

(H) Performance metrics in the validation cohort for each DNA methylation class for samples belonging to MNP group 2.

(I) Performance metrics in the validation cohort for each DNA methylation class for samples belonging to MNP group 3.

(J) Confusion matrix showing the misclassified samples at DNA methylation class level for the validation data when MNP v11b classification was used as the ground truth. The majority of the misclassified samples are glioblastoma multiformes (GBMs) and fall into the other GBM, CONTROL INFLAM, or CONTROL REACT classes. GBM MES tumors were also misclassified as low-grade gliomas, and 10 different tumor samples as control tissue from pituitary gland anterior lobe (CONTR ADENOPIT). The majority of the misclassified samples belong to low-confidence MNP Groups 2-3.

(K) Confusion matrix showing the misclassified samples at diagnostic tumor class level for the validation data when MNP v11b classification was used as the ground truth.

**Supplementary Figure S4**

Calibration curve for the SVM confidence score (Diff12/Prob1) in the external validation cohort. The plot shows the relationship between the confidence score and the observed classification accuracy across confidence score bins. Higher confidence scores were consistently associated with higher observed classification accuracy, indicating that the confidence score provided meaningful stratification of prediction reliability.

**Supplementary Figure S5**

(A) The count of captured features out of the 1003 intended per sample in targeted EM-seq data using IDT xGen Hybridization Core Reagents v2

(B) The coverage of captured features per sample in targeted EM-seq data using IDT xGen Hybridization Core Reagents v2

(C) The count of captured features out of the 1003 intended per sample in targeted EM-seq data using IDT xGen Hybridization Core Reagents v3

(D) The coverage of captured features per sample in targeted EM-seq data using IDT xGen Hybridization Core Reagents v3

**Supplementary Figure S6**

Median absolute error of imputed beta values of the selected 1003 features across an increasing proportion of randomly removed values. The x-axis shows the percentage of missing values measured between 10% and 90%. For each level of missingness, values were randomly removed and imputed 10 times. The y-axis shows the median absolute error of predicted beta values measured from the 10 folds. Blue line denotes median while lower and upper limits of the light blue shaded area indicates the first and third quartiles of absolute error. Results are shown for both the validation cohort (GSE90496), imputed against Capper et al. (2018) reference cohort, and the enzymatic methylation sequencing (EM-seq) batch 1, carried out with IDT xGen Hybridization Core Reagents v2, imputed against a combined Capper *et al.* (2018) reference and validation cohort. Median absolute error increased slightly along with an increased number of missing values. When 50% of values were masked, they were predicted with 0.064 (IQR 0.028-0.158) and 0.061 (IQR 0.028-0.149) for the nanopore and EM-sequencing cohorts, respectively. Only the first EM-sequencing batch was chosen as it had near perfect coverage of the targeted 1003 CpGs.

**Supplementary Figure S7**

(A) The count of captured features out of the 1003 intended per sample in SureSelect data

(B) The coverage of captured features per sample in SureSelect data

(C) The count of captured features out of the 1003 intended per sample in cfRRBS data

(D) The coverage of captured features per sample in cfRRBS data

(E) The count of captured features out of the 1003 selected CpG sites per sample in the Nanopore sequencing data

(F) The mean coverage of captured features per sample in the Nanopore sequencing data

**Supplementary Figure S8**

Methylation analysis at the MGMT promoter region in batch 1 (A) and batch 2 (B) of EM-seq samples. Dark grey shading indicates beta values (0–1) while blue shading the sequencing depth (for batch 1 0–2,728 and for batch 2, 0–100 (maximum depth 350)). The dashed grey line indicates 0.3 beta value, a commonly used threshold for a methylated MGMT promoter. Brown and green bars indicate EM-seq target-enrichment probes across the MGMT promoter region relative to genomic coordinates (chr10:131,264,949). The locations of diagnostically relevant Differentially Methylated Region 1 (DMR1), DMR2, and two commonly utilized CpGs are highlighted. All oligodendrogliomas show the characteristic pattern of MGMT promoter methylation.

**Supplementary Figure S9**

Reference-free copy number analysis from off-target reads of targeted enzymatic sequencing (EM-seq) using 500 kb binning. Y-axis shows the estimated copy number ratio on a log2 scale relative to its genomic coordinates. From all oligodendrogliomas, glioblastomas, and meningiomas, their characteristic alterations can be observed.

**Supplementary Figure S10**

Copy number analysis comparing 5 samples, S23-S27 (subpanels A-E, respectively), for which both EM-seq (left) and WGS (right) data was available, show the profiles to have a near perfect concordance. Despite differences in absolute copy-number values, the same underlying alterations are represented on different absolute scales. WGS copy numbers were obtained from Ohlsbom S. et al. Multi￼region whole￼genome and transcriptomic profiling uncovers plastic, subclone￼linked cell states in high￼grade diffuse astrocytomas. bioRxiv. doi:10.64898/2026.08.11.743949, derived using Battenberg v2.2.10 algorithm, which estimates tumor purity and ploidy while using matched normal data as a reference. In QDNAseq (v1.42.0) -packages algorithm (used for EM-seq samples), the most common copy-number state is assumed to be diploid, due to the lack of a reference.

**Supplementary Figure S11**

Copy number analysis of the CDKN2A locus based on targeted sequencing sites (5 probes total). (A) Boxplot showing tumor purity-adjusted log2 ratios of normalized read counts at targeted CDKN2A exon sites in EM-seq tumor samples from the first sequencing batch (S01-S15, S19, and S20). Log2 ratios were calculated relative to read depths in matched whole blood cell (WBC) control samples from the same batch (S16-S18). Tumor fraction adjustment was not applied to samples without an estimated tumor fraction (S23-S35, S40-S43, and S57). (B) Tumor purity-adjusted log2 ratios plotted across genomic coordinates spanning the CDKN2A locus (hg19 reference genome).

**Supplementary Figure S12.**

(A) Distance from the 1003 features (probe index CpGs) to the nearest CpG measured by EM-seq.

(B) Distance from the 1003 features (probe index CpGs) to the nearest CpG measured by SureSelect.

(C) Distance from the 1003 features (probe index CpGs) to the nearest CpG measured by cfRRBS.

**Supplementary Figure S13**

(A) The distribution of SVM confidence scores and misclassified samples is retained through the reduction of features.

**Supplementary Figure S14**

Sample-wise DNA methylation values of 298 select features, represented as beta values (range 0-1), in the reference cohort. Samples have been categorized based on their DNA methylation class and Diagnostic class labels. Features that are also part of the 163-feature set are annotated on the left side of the heatmap.

**Supplementary Figure S15**

Sample-wise DNA methylation values of 298 select features, represented as beta values (range 0-1), in the validation cohort. Samples have been categorized based on their DNA methylation class and Diagnostic class labels. Features that are also part of the 163-feature set are annotated on the left side of the heatmap.

**Supplementary Figure S16**

Diagnostically relevant class named “Medulloblastoma, WNT-activated” (MB, WNT) can be separated with perfect precision from the reference embryonal tumor classes using only two features for each comparison. Distinguishing the MB, WNT using probes cg05908105 (beta ≤ 0.3, or beta ≤ 0.5) and cg08123444 (beta ≤ 0.5) in an independent cohort containing 52 MB, WNT cases and 253 other MBs (accuracy: 0.974 [CI95; 0.949-0.987], F1 score: 0.917 [CI95; 0.815-0.968], precision: 1.000 [CI95; 0.920-1.000], recall: 0.846 [CI95; 0.725-0.920]). Ninety-five percent confidence intervals (in square-brackets) for accuracy, precision, and recall were calculated using Wilson score intervals, and confidence intervals for the F1 score were estimated by nonparametric bootstrap resampling (100,000 iterations). TP: True positive, FN: False negative, FP: False positive, TN: True negative.

**Supplementary Table S1.**

The full and abbreviated class terms for DNA methylation classes and diagnostic classes.

**Supplementary Table S2.**

DNA methylation classes and ENLR coefficient values across tumor methylation classes. “FeatSetCoord”-sheet contains the consistency class of each feature included in the 4976 feature set or its subsets across DNA methylation classes, where: 1 = consistently unmethylated, 2 = consistently intermediately methylated, 3 = consistently methylated, 0 = inconsistent. It also provides the genomic coordinates of each feature and indicates the reduced probe set(s) to which each feature belongs, as well as, presents how the probes selected with either multi- or inter-class consistency approaches are retained throughout the feature reductions (at the bottom of the sheet). The MetValDifAnalysis table contains the best performing probes for finding consistent group-specific features for separating selected tumor types from each other. The Coefs1003Feats91Classes-sheet contains ENLR coefficient values for the select 1003 features across tumor methylation classes and Classwise_Jaccard_Index contains classwise ENLR feature selection stability based on Jaccard similarity and coefficient variability. DifferingConsFeatures_1003 and DifferingConsFeatures2vs13_1003 report the number of differing consistent features in pair-wise DNA methylation group comparisons.

**Supplementary Table S3.**

Gene set enrichment analysis of genes to which the selected set of features (1003) were annotated. The results for analyses done considering either all the genomic locations with respect to the gene, all the locations except for the promoter or only the promoter are in separate sheets.

**Supplementary Table S4.**

Results of the SVM and MNP classification with 1003 features in the external validation cohort of public microarray datasets (total of 1993 samples, representing 72 distinct tumors and four control methylation classes), showing Capper (1094 samples) and GEO (899 samples) validation cohorts in separate sheets. Presented are the SVM predicted labels in comparison to MNP labels in both v11b and v12.8 models as well classification consistency based on diagnosis-assisted evaluation. For comparison, both MNP and SVM confidence scores and groups for each sample are presented. Furthermore, sample counts and classification performance in the high confidence SVM group with different confidence cutoffs is shown in ‘SVM confidence cutoff comp’ sheet.

**Supplementary Table S5.**

DNA methylation sequencing cohort. The table presents for each sample: Patient ID, Pathological diagnosis, Tumor grade, IDH mutation status, Age category at surgery, Sex, Disease stage, Material type, experienced pathologist’s estimate of tumor purity (Estimated tumor purity %) based on histological analysis, and whether the sample was analyzed with EM-seq (EM), SureSelect (SS), or cfRRBS (CF) sequencing (Sequencing method).

**Supplementary Table S6**.

The measured feature-wise CpG beta values with and without imputed values from sequenced samples in EM-seq, SureSelect, cfRRBS, and Nanopore sequencing datasets.

**Supplementary Table S7**.

Summary of classification results (1^st^, 2^nd^ and 3^rd^ predictions with associated SVM confidence scores) for all the sequencing samples analyzed with targeted sequencing approaches and classified using our classifier with either v11b labeling or v12.8 labeling. In addition, we report the predictions of our classifier using external Nanopore-sequencing data (n=208) with v11b labels.

**Supplementary Table S8.**

The ”MGMT_promoter_analysis”-sheet contains measured beta values and coverages of MGMT promoter relevant CpGs #31 [CpG 31: chr10:131265209, hg19] and #84 [CpG 84: chr10:131265575, hg19], along with defined DMR1 [Differentially methylated region 1 (DMR1): CpGs 25-50, chr10:131265158-131265340, hg19] and DMR2 [Differentially methylated region 2 (DMR2): CpGs 73-90, chr10:131265496-131265626, hg19] regions for all sequenced samples.

**Supplementary Table S9.**

SVM confidence scores and calculated beta values of features for the samples analyzed with EM-seq (batch 1), Sureselect or cfRRBS when imputing was not used. Beta values were calculated by including the closest CpG from the index CpG with enough coverage within maximum 200 bp from the index CpG and expanding the region 50 bp to both directions from this closest CpG and calculating the mean methylation of CpGs within that region.

**Supplementary Table S10.**

The SVM classification results of the 1993 samples when 1003, 533, 298, or 163 features were used for the classification. Both the predicted class labels (pred) and SVM confidence scores are shown for each sample (SampleID).
