## Supplementary figures and images for "Consistent DNA methylation patterns enable accurate and interpretable cross-platform classification of central nervous system tumors"

### Figure S1

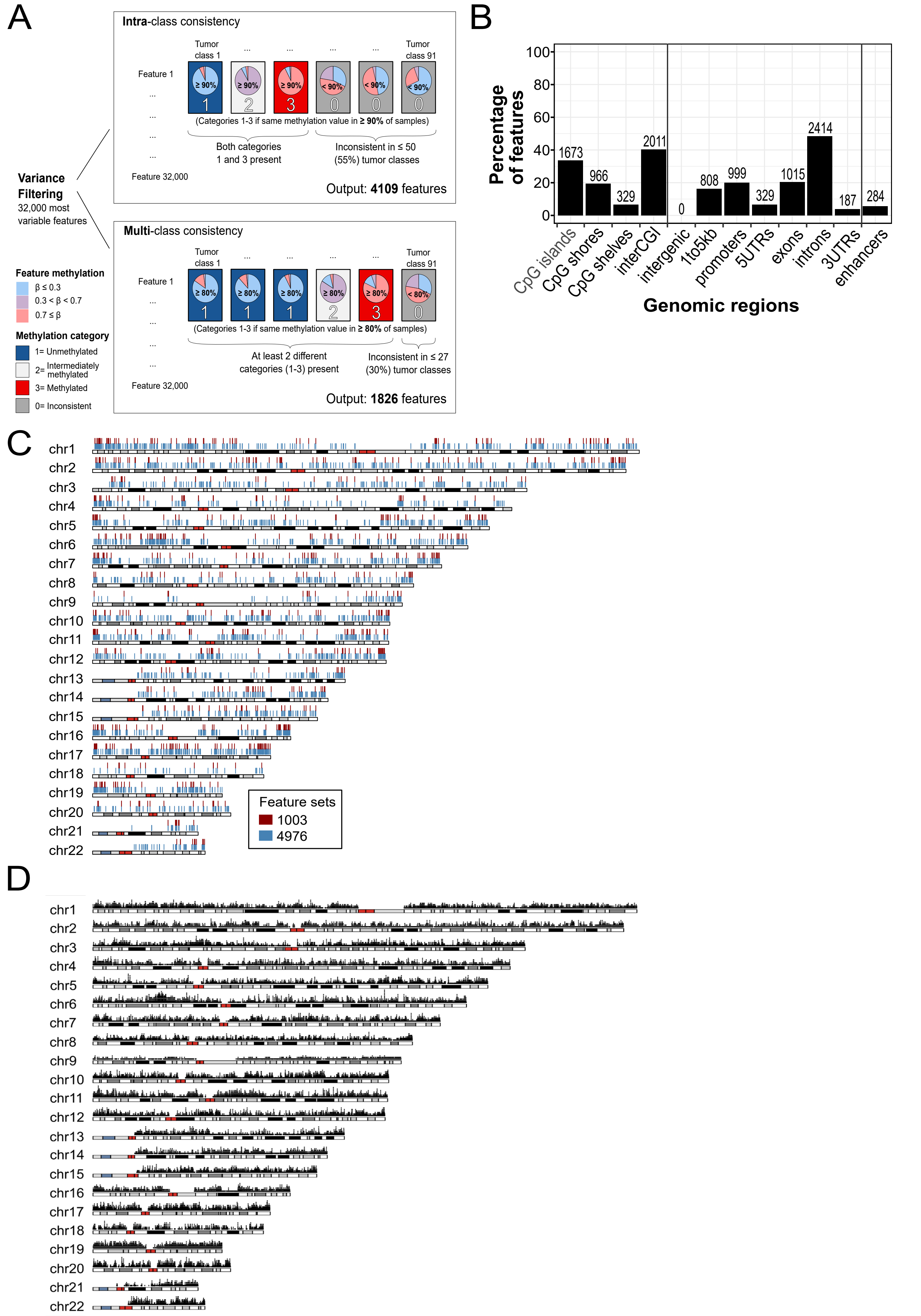

### Figure S2

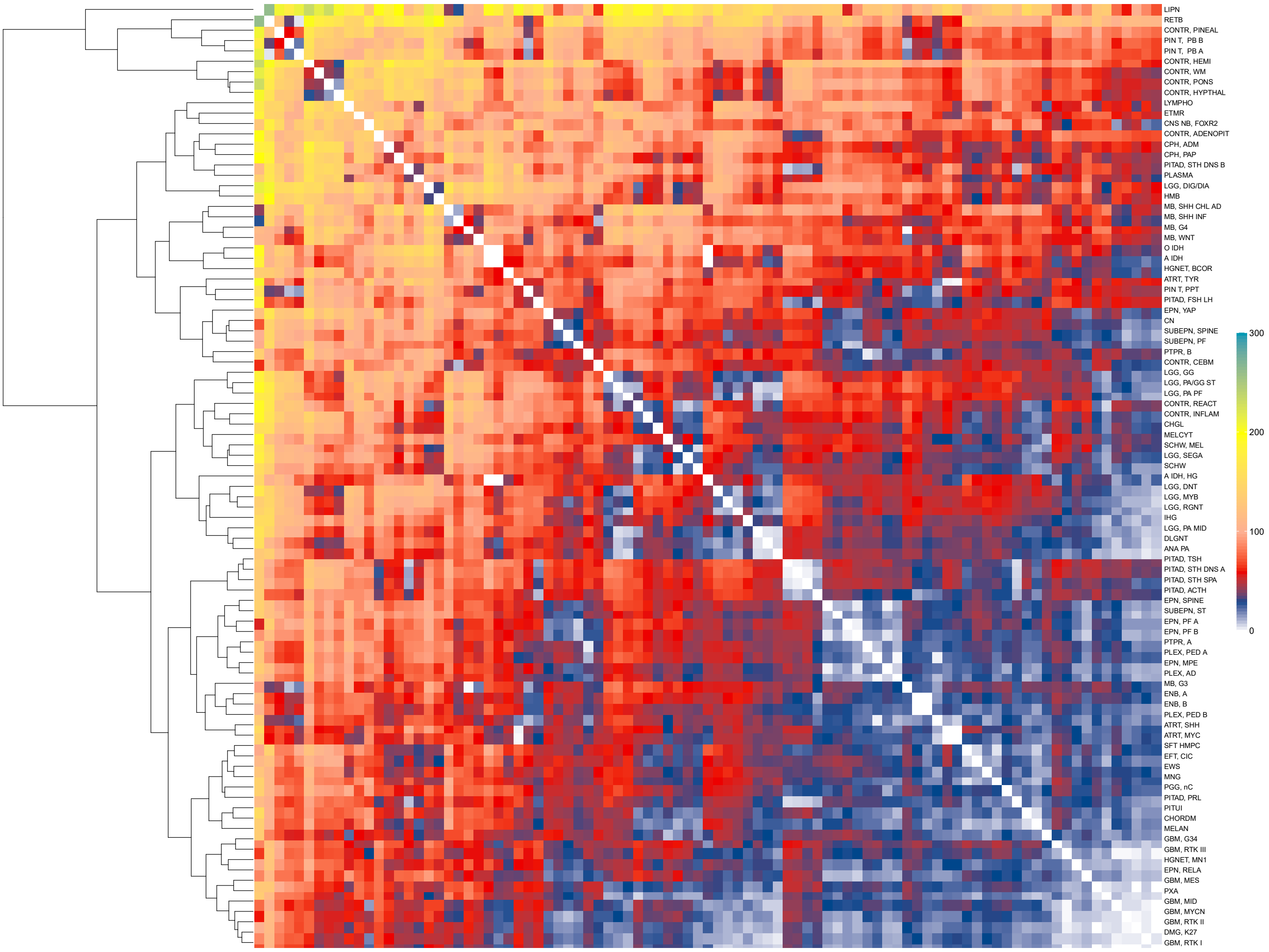

### Figure S3

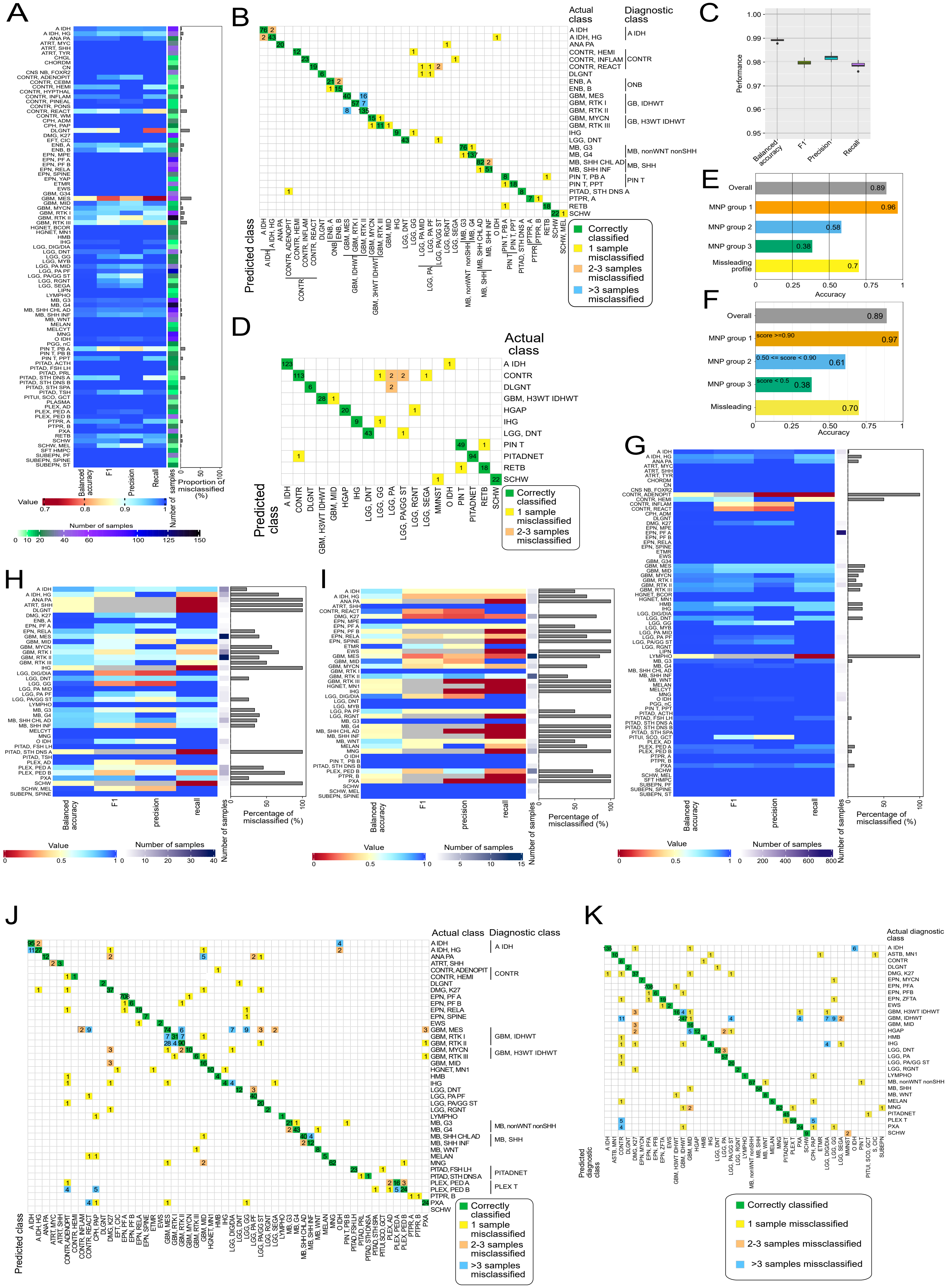

### Figure S4

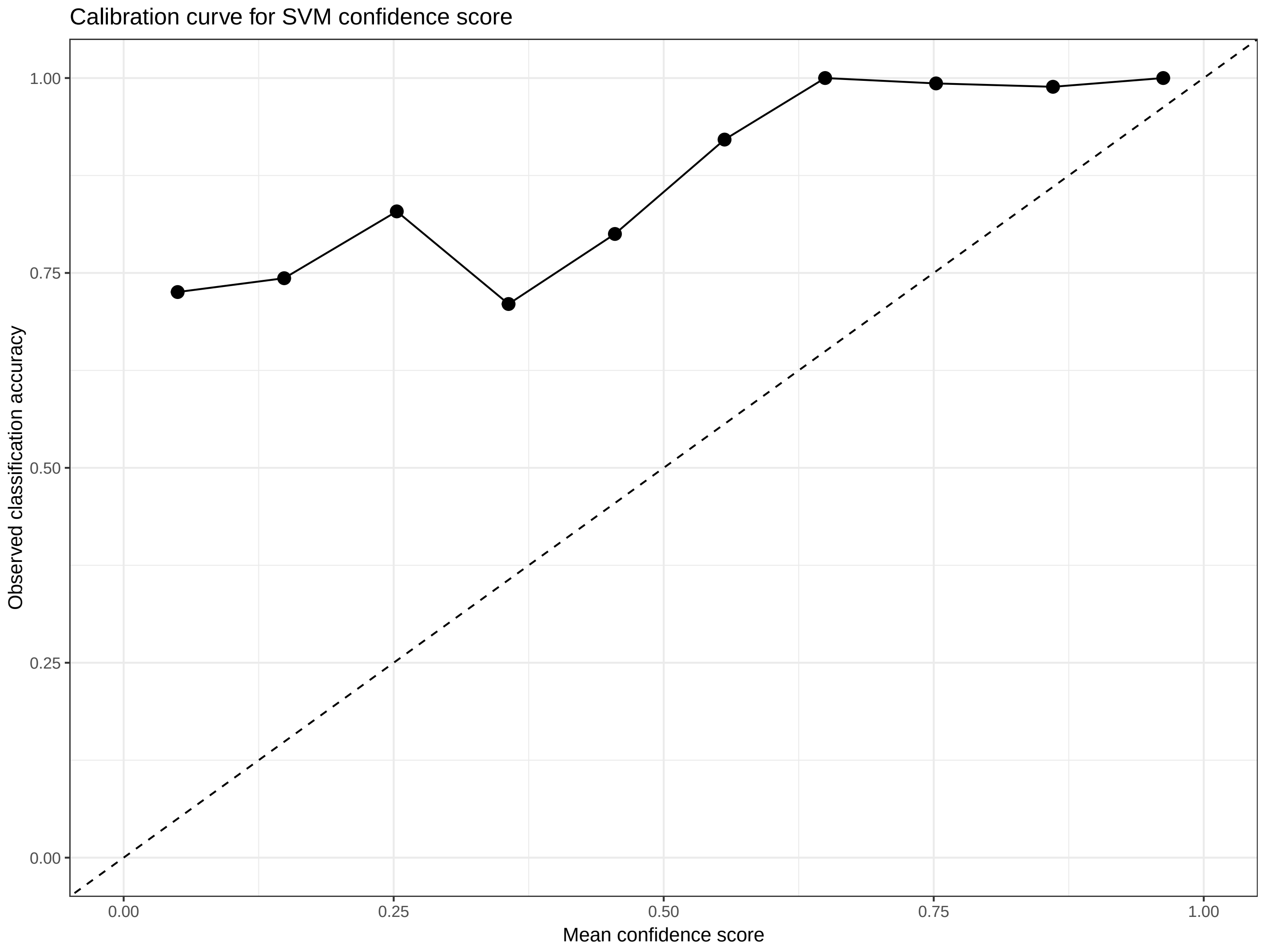

### Figure S5

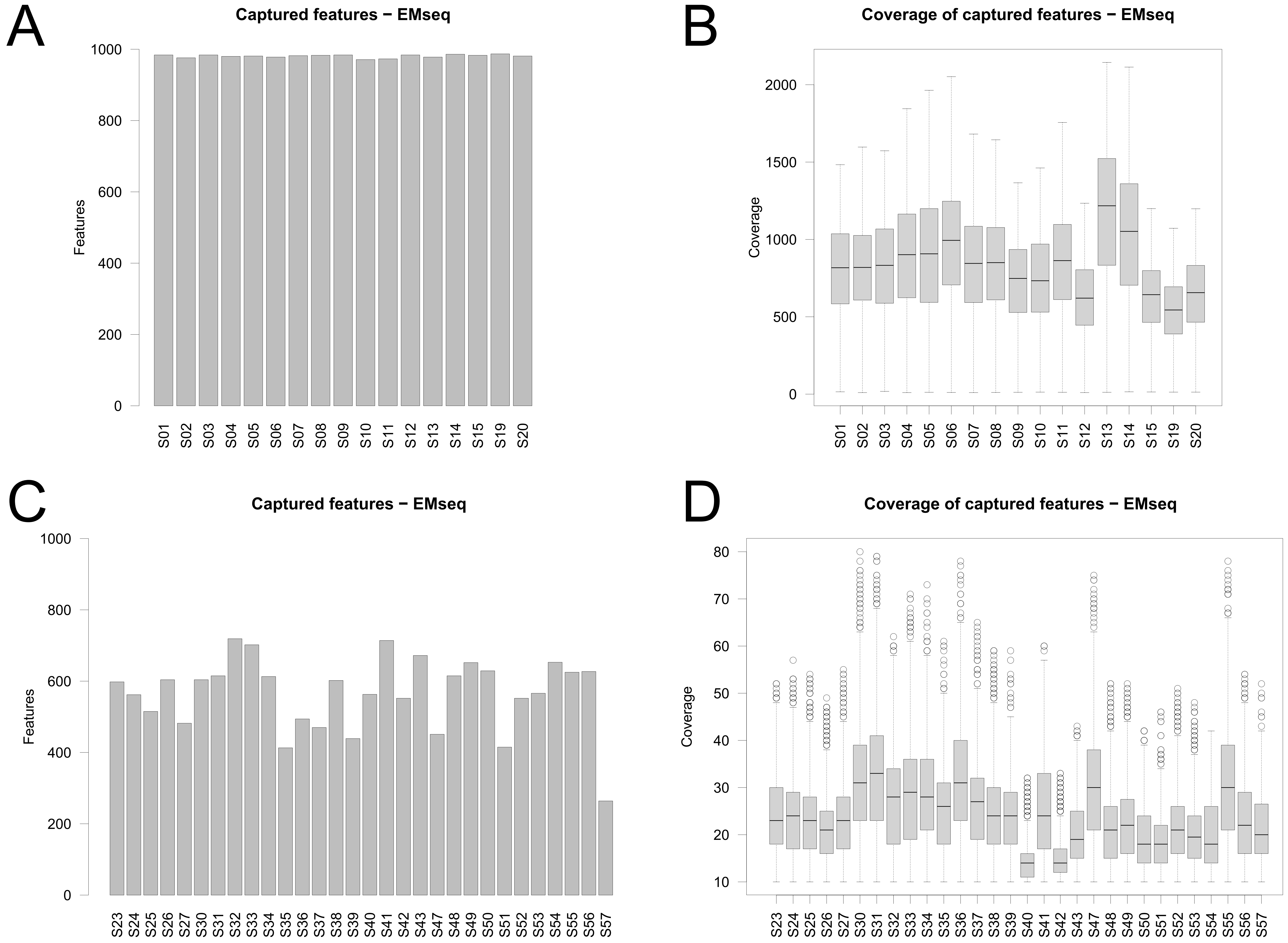

### Figure S6

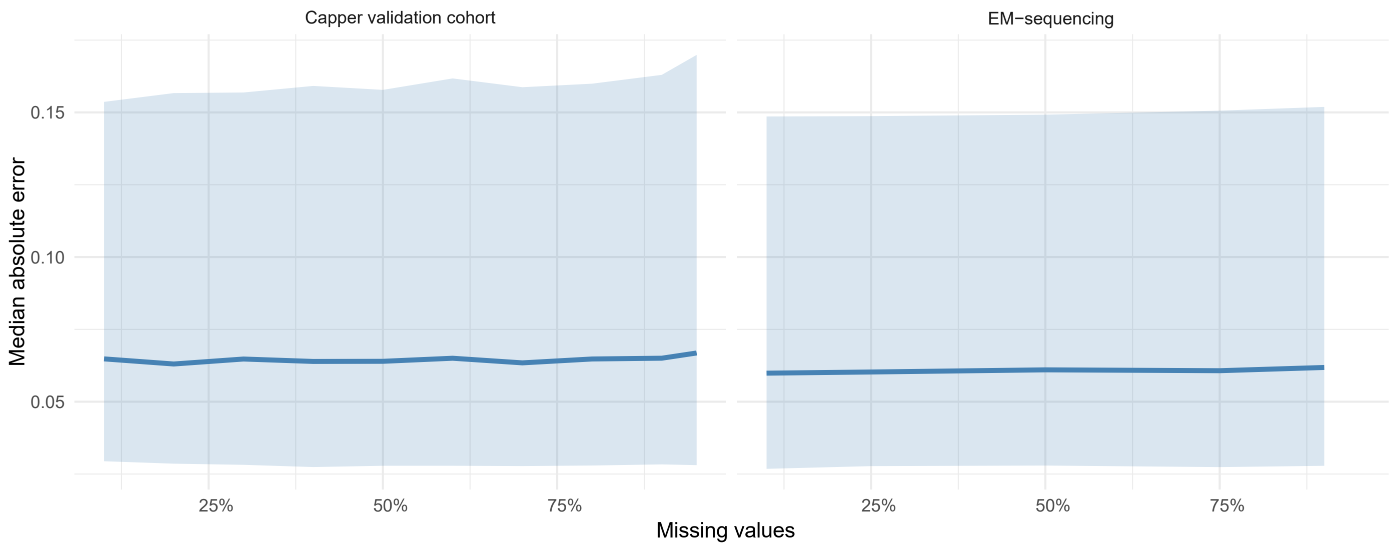

### Figure S7

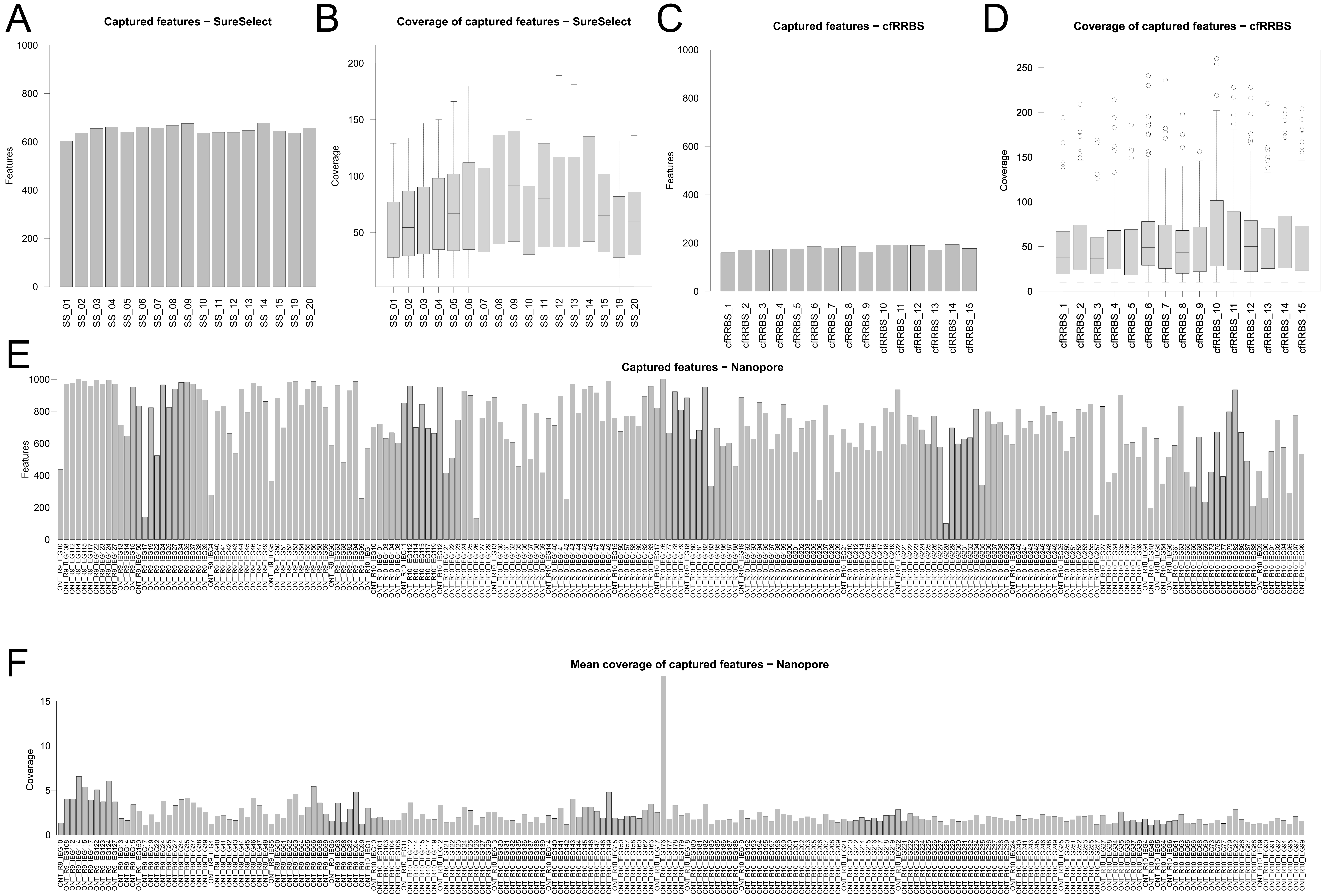

### Figure S8

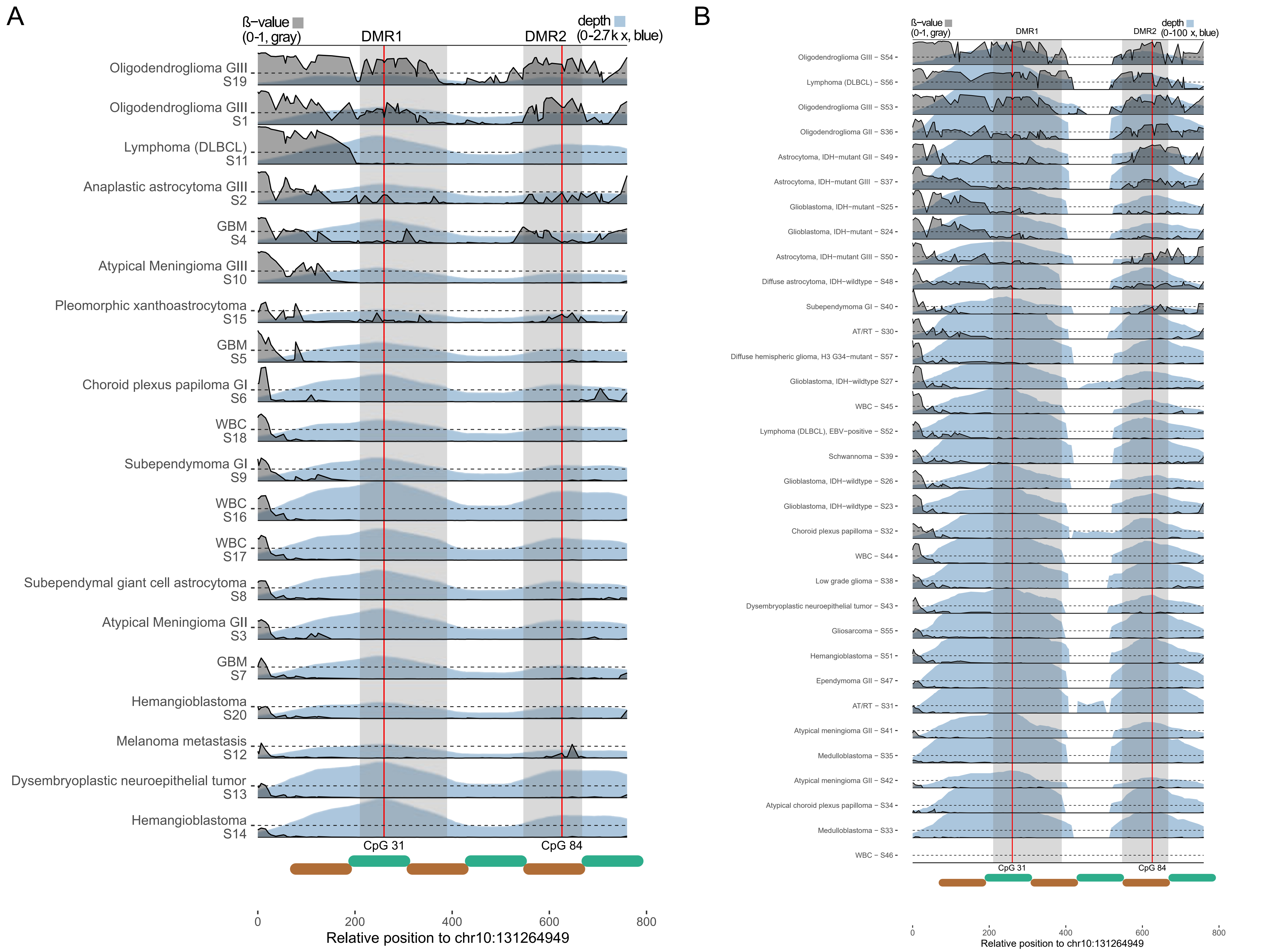

### Figure S9

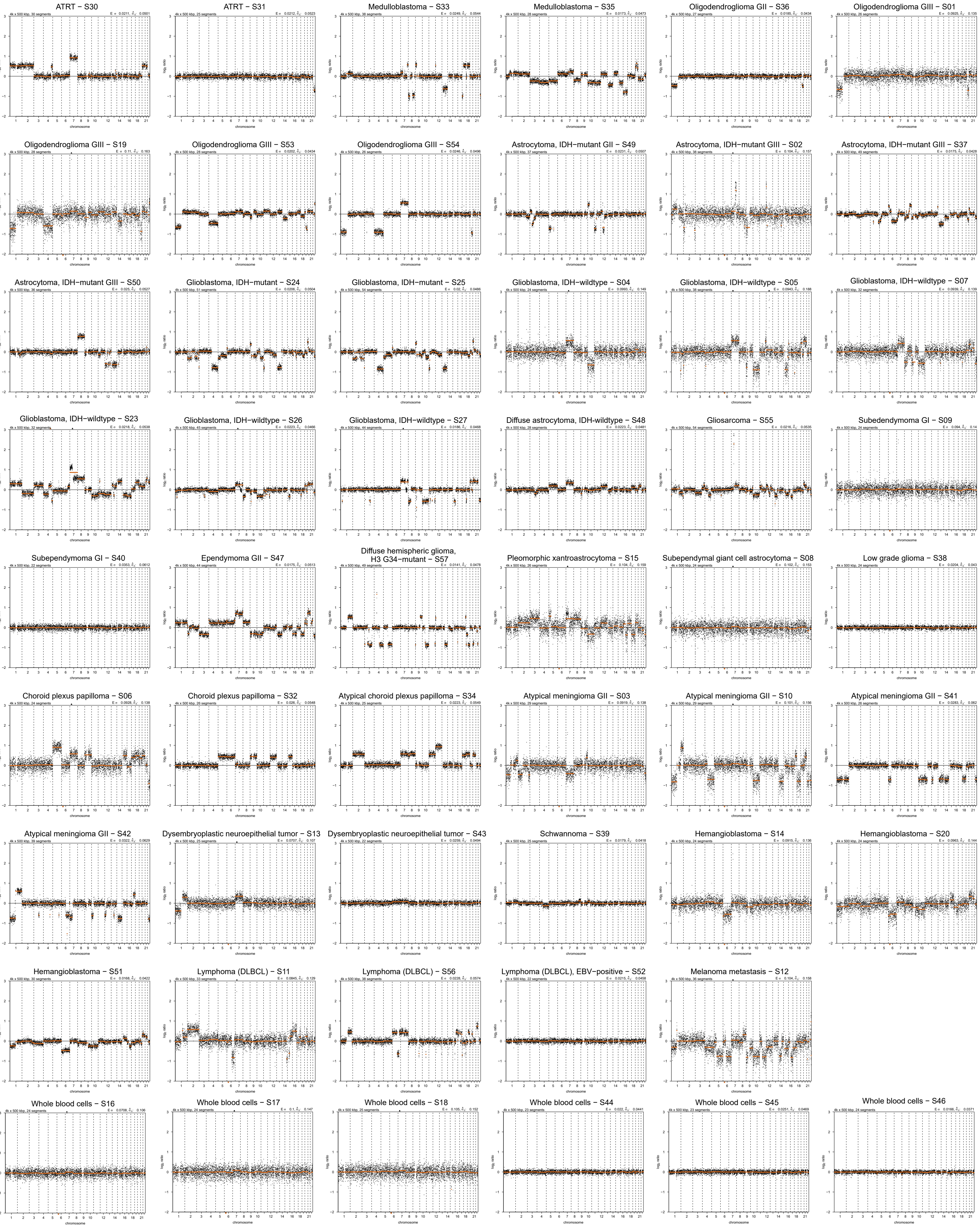

### Figure S10

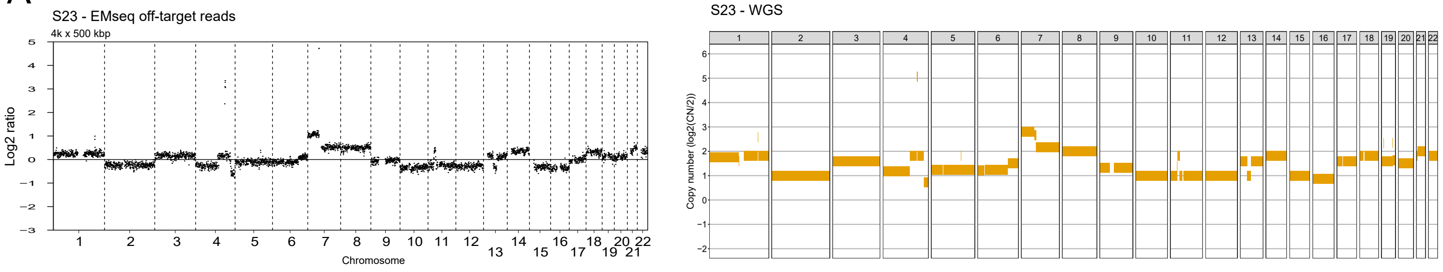

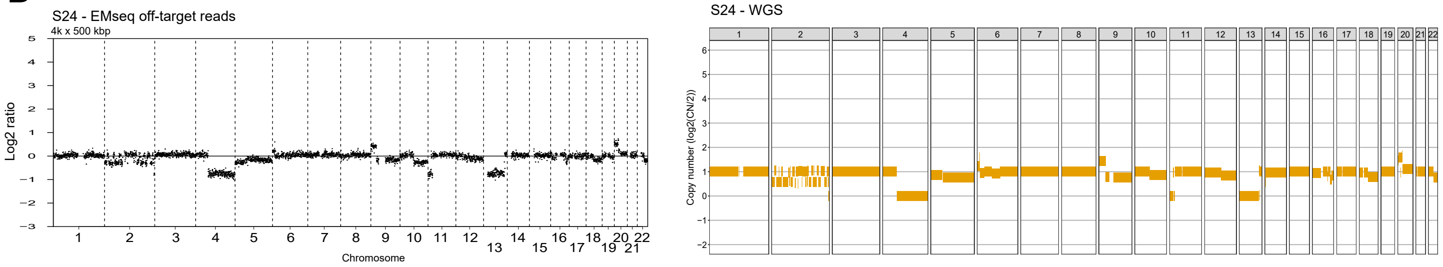

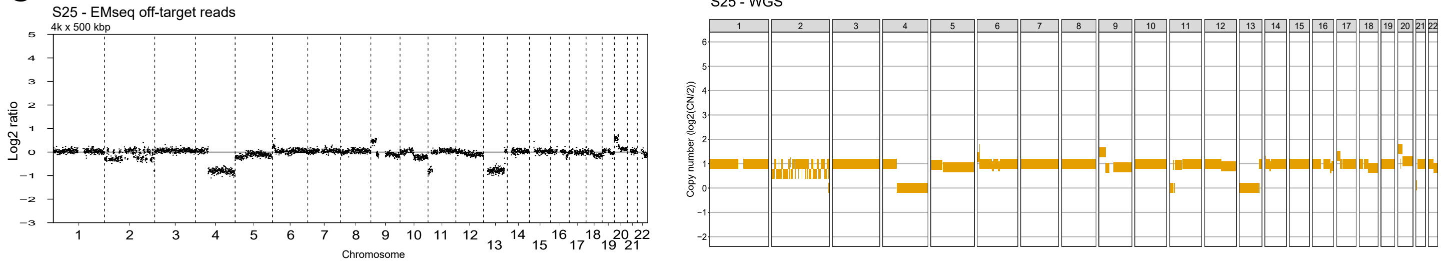

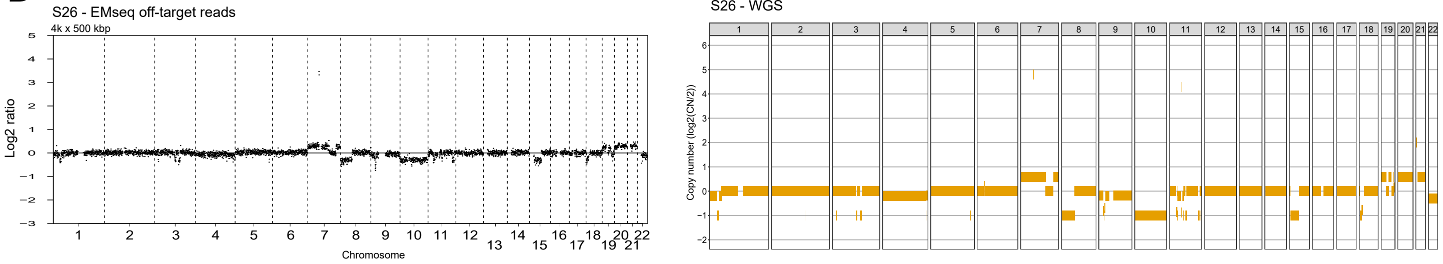

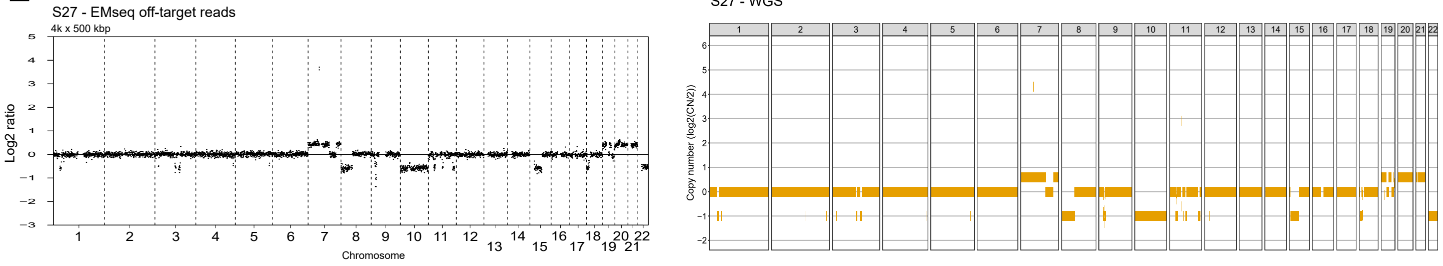

### Figure S11

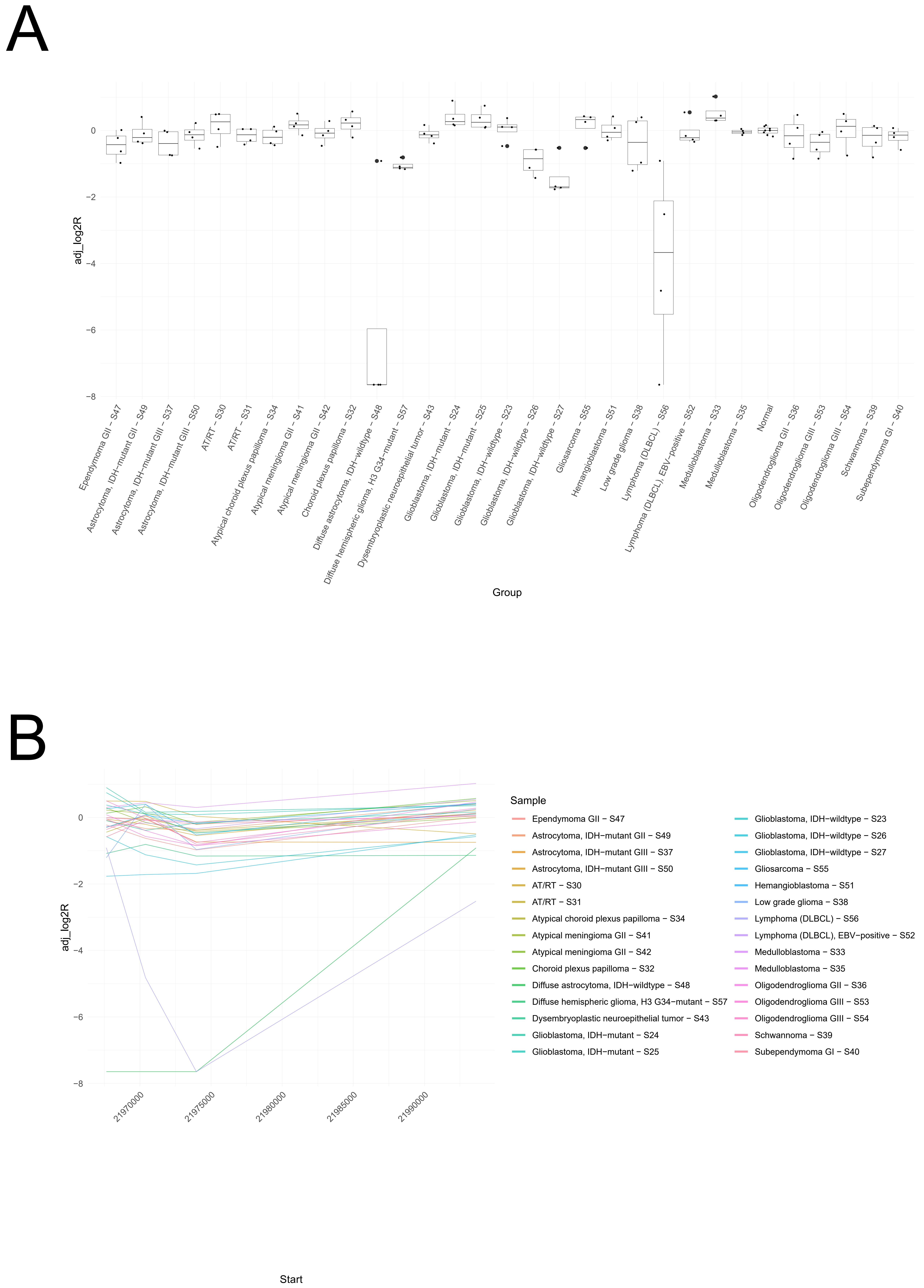

### Figure S12

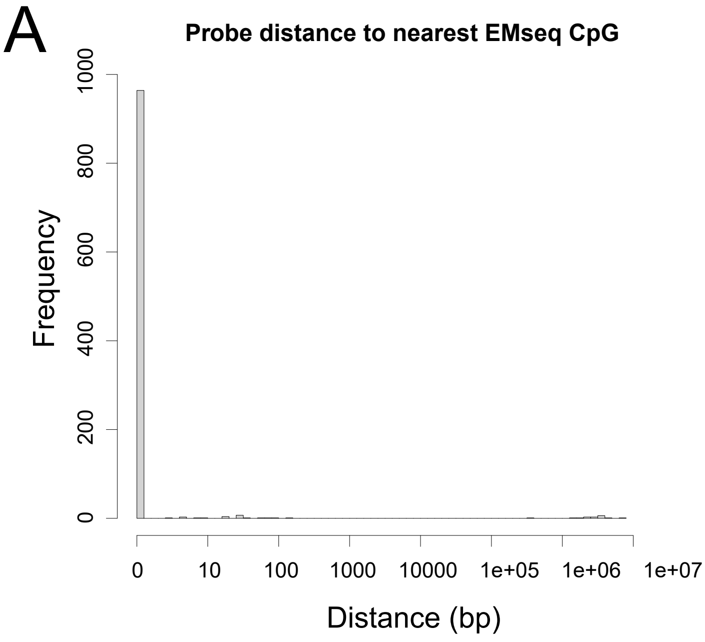

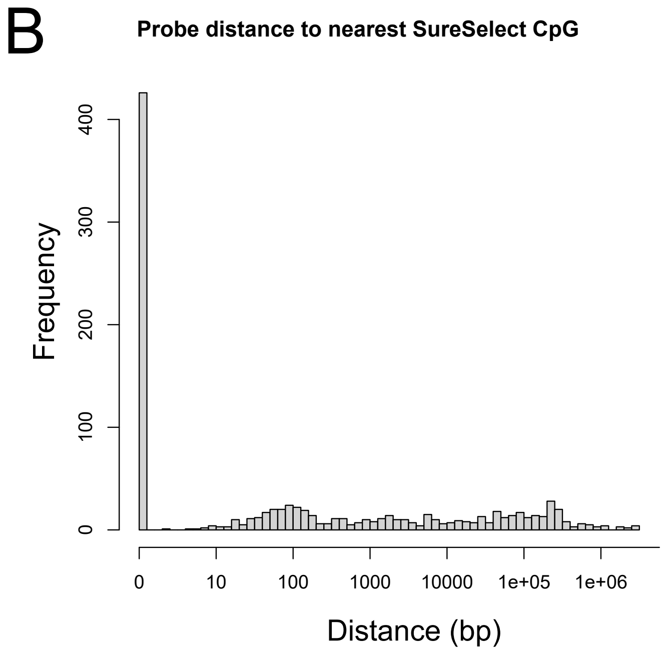

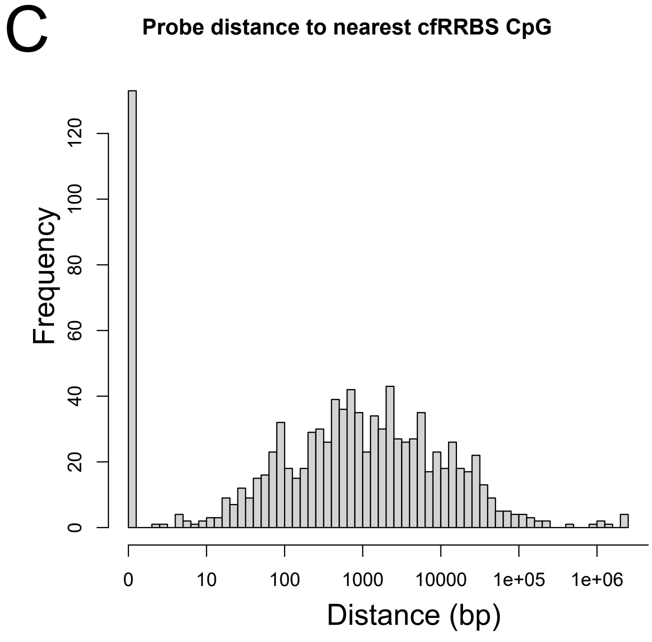

### Figure S13

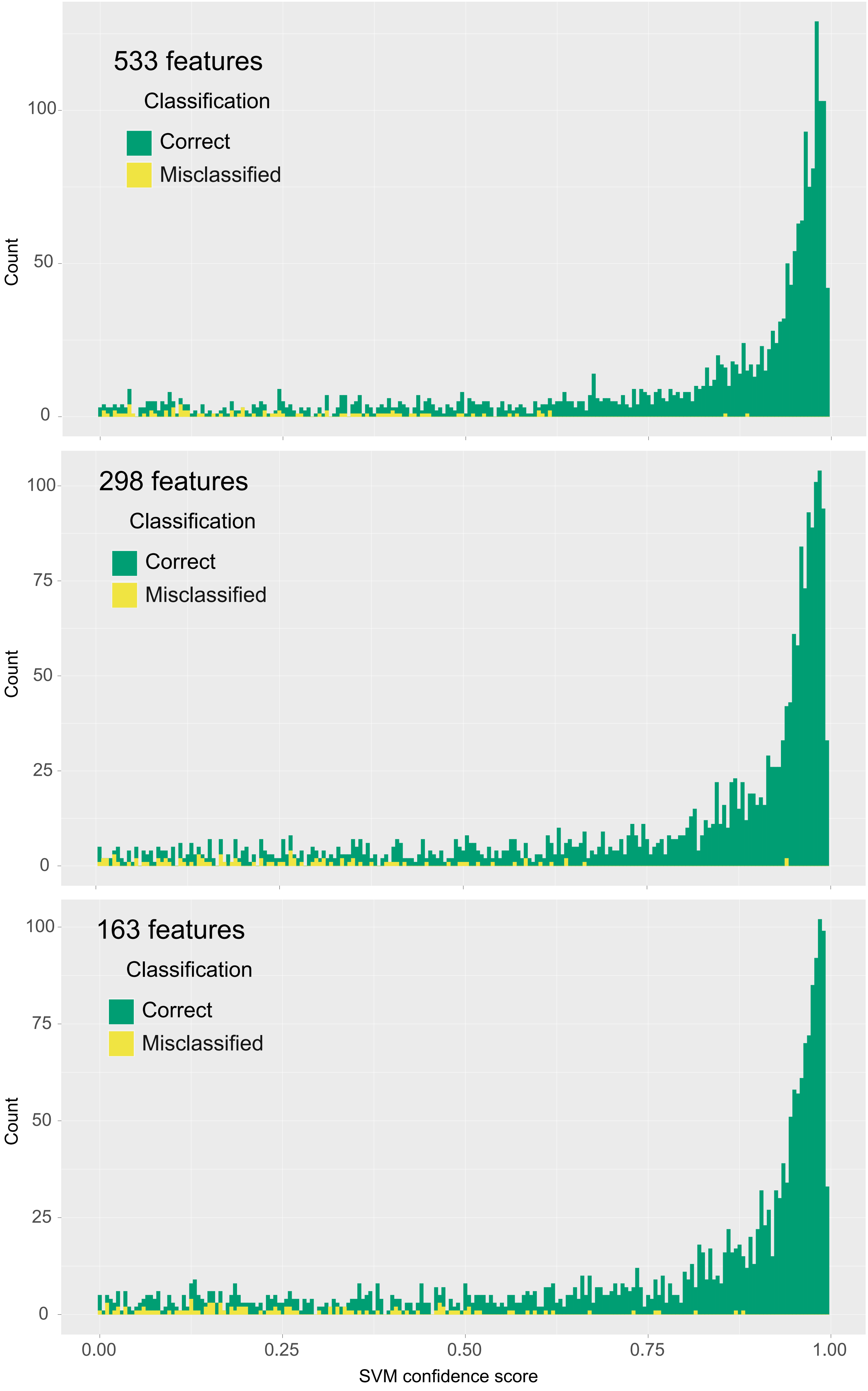

### Figure S14

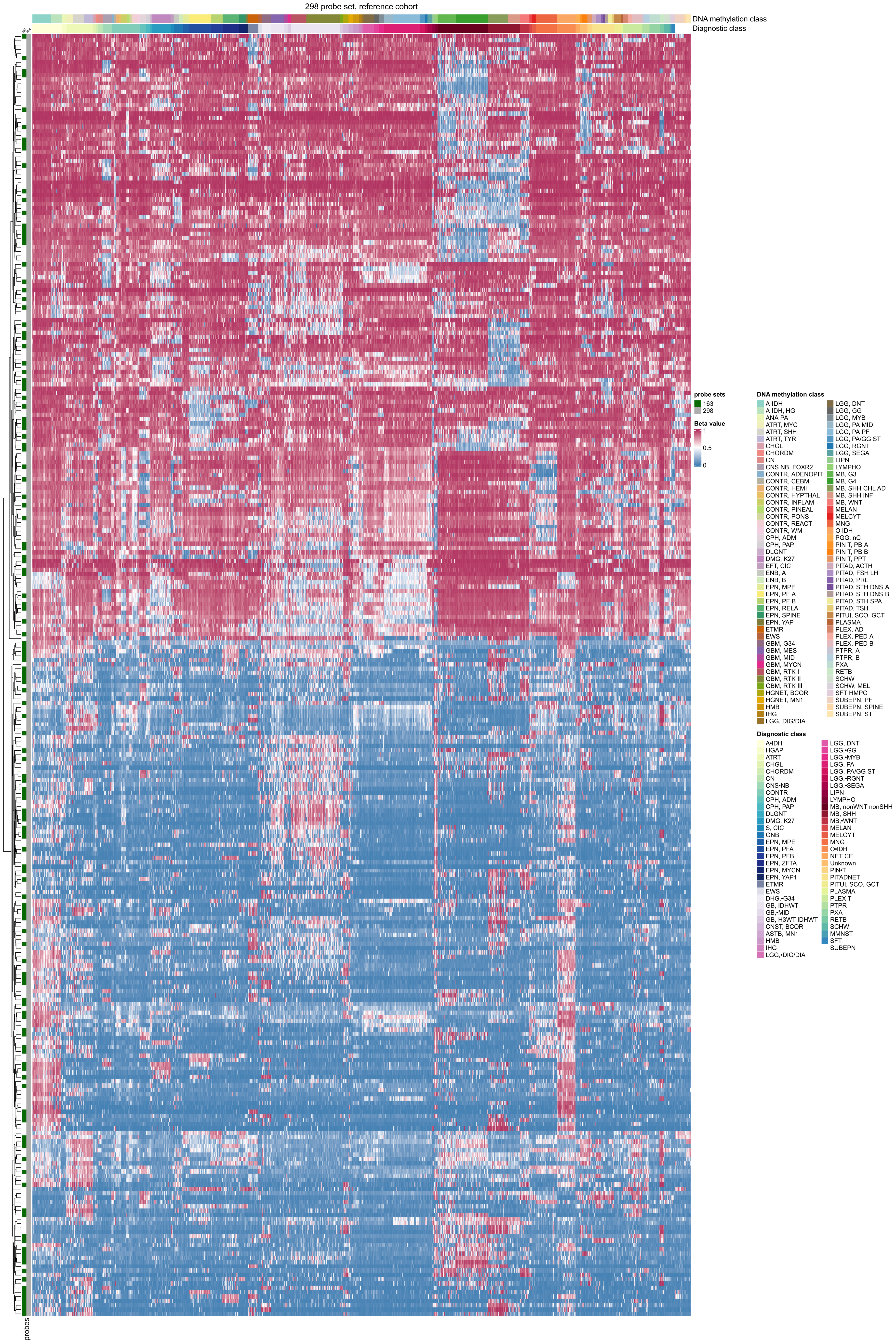

### Figure S15

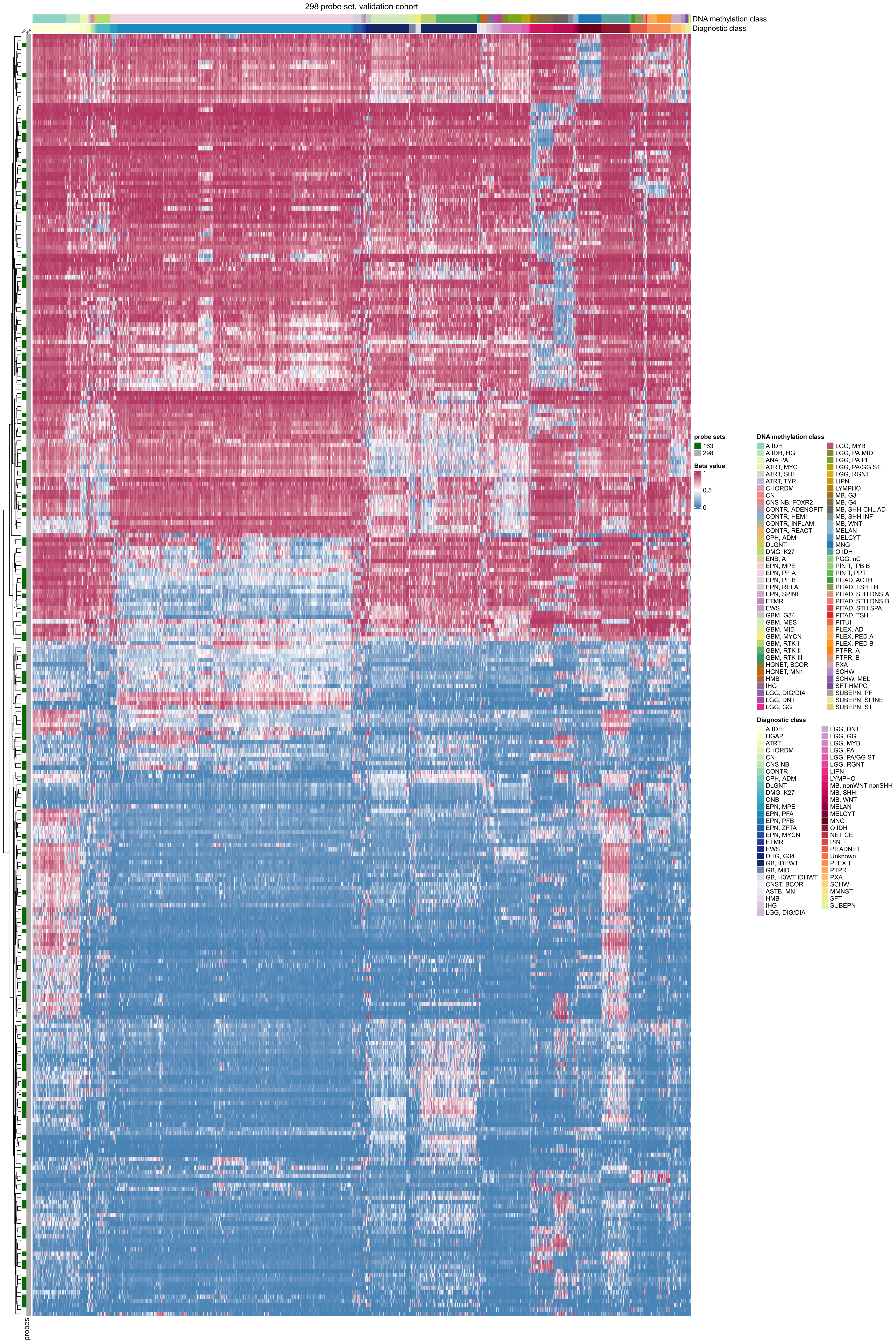

### Figure S16

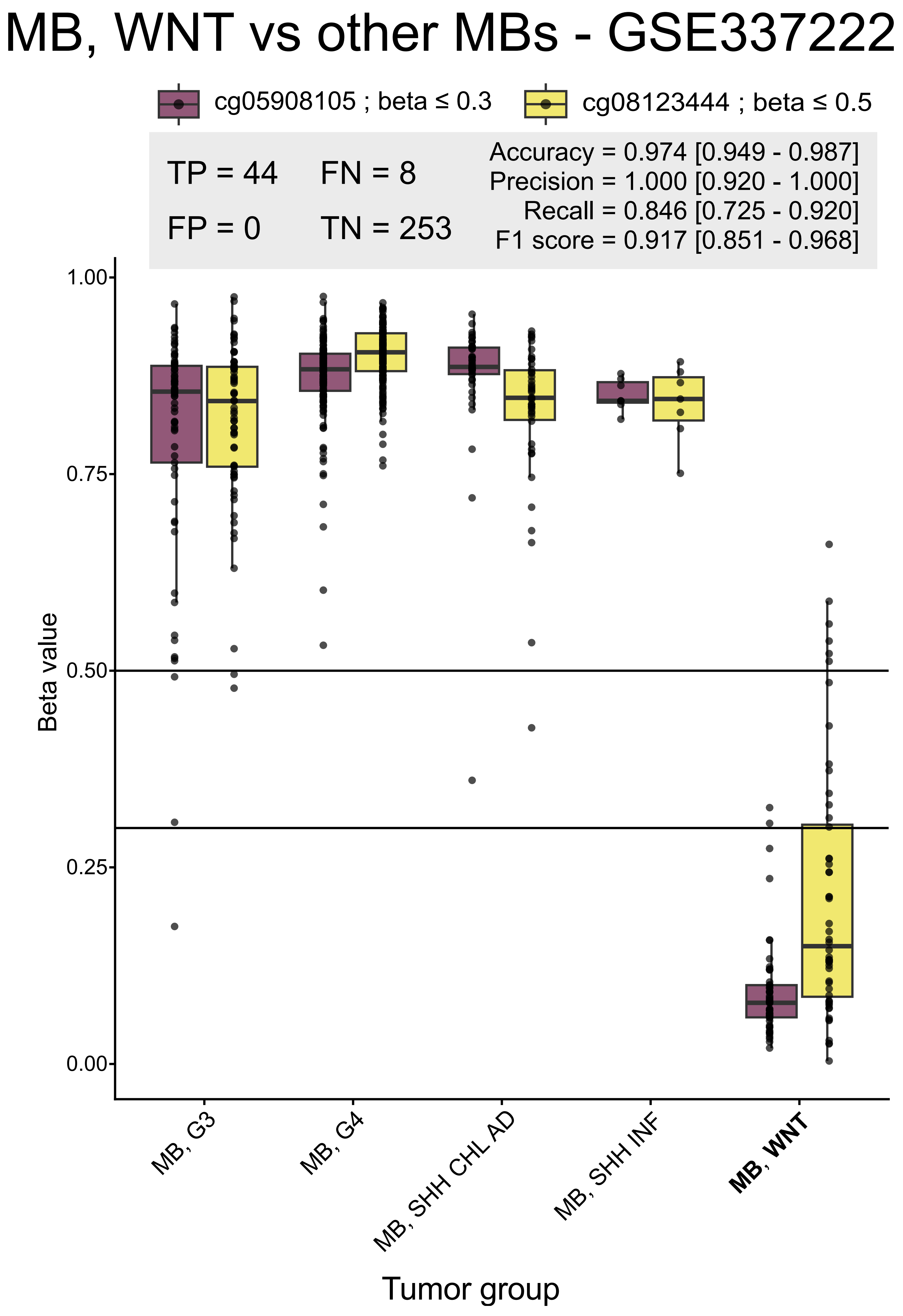
